# Multi-organ imaging-derived polygenic indexes for brain and body health

**DOI:** 10.1101/2023.04.18.23288769

**Authors:** Xiaochen Yang, Patrick F. Sullivan, Bingxuan Li, Zirui Fan, Dezheng Ding, Juan Shu, Yuxin Guo, Peristera Paschou, Jingxuan Bao, Li Shen, Marylyn D. Ritchie, Gideon Nave, Michael L. Platt, Tengfei Li, Hongtu Zhu, Bingxin Zhao

**Author notes:** Corresponding authors*: Xiaochen Yang, Hongtu Zhu, and Bingxin Zhao. Lead contact:* Bingxin Zhao 413 Academic Research Building 265 South 37th Street, Philadelphia, PA 19104.

## Abstract

The UK Biobank (UKB) imaging project is a crucial resource for biomedical research, but is limited to 100,000 participants due to cost and accessibility barriers. Here we used genetic data to predict heritable imaging-derived phenotypes (IDPs) for a larger cohort. We developed and evaluated 4,375 IDP genetic scores (IGS) derived from UKB brain and body images. When applied to UKB participants who were not imaged, IGS revealed links to numerous phenotypes and stratified participants at increased risk for both brain and somatic diseases. For example, IGS identified individuals at higher risk for Alzheimer’s disease and multiple sclerosis, offering additional insights beyond traditional polygenic risk scores of these diseases. When applied to independent external cohorts, IGS also stratified those at high disease risk in the All of Us Research Program and the Alzheimer’s Disease Neuroimaging Initiative study. Our results demonstrate that, while the UKB imaging cohort is largely healthy and may not be the most enriched for disease risk management, it holds immense potential for stratifying the risk of various brain and body diseases in broader external genetic cohorts.

## Introduction

The large-scale imaging data from the UK Biobank (UKB) imaging study has proven to be an immensely valuable resource for characterizing the structural and functional organizations of the human brain and body^1–4^. These data have been used to establish links with clinical biomarkers^5,6^, predict biological aging^7–9^, study socioeconomic outcomes^10,11^, and facilitate early disease detection^12^. Launched in 2014, the UKB imaging project reached a milestone in 2022 by completing the multimodal brain magnetic resonance imaging (MRI) of its 60,000th participant. Although it is already the world’s largest imaging study, the UKB imaging study will ultimately include nearly 100,000 participants^4^, or about 20% of all UKB participants. Given the cost and difficulty of collecting additional imaging data, it is crucial to develop strategies that extend the utility of the existing UKB imaging data.

Polygenic indices^13^ (also known as genetic scores, polygenic scores, or polygenic risk scores [PRS]) can be used to predict traits or disease genetic risk by aggregating genetic information across the genome^14–21^. The development of numerous prediction methods^22^, reporting standards^23^, genetic data resources^24^, and data sharing platforms^25^ has enabled the application of these indices to a wide variety of complex diseases and traits. Both family and population-based studies have shown that variations in brain and body, as measured by structural and functional MRI, are heritable^2,26–32^. Recent genome-wide association studies (GWAS)^2,29,33–41^ have identified many genetic loci associated with brain and body imaging-derived phenotypes (IDPs). Given the cost and complexity of imaging phenotypes, it is natural to ask whether genetic data can be used to predict IDPs for UKB participants who are not imaged or who are part of other cohorts of interest (e.g., another biobank-scale cohort, or a cohort dedicated to Alzheimer’s disease). Several GWAS have reported the prediction accuracy (out-of-sample *R*^2^) of genetically predicted brain IDPs^33,42,43^ in small-scale independent testing data, indicating that genetic predictors could partially recover imaging variations. These pilot studies demonstrate that IDP genetic scores (IGS) can serve as valuable proxy biomarkers of the brain and body in the absence of readily available imaging data^29,40,44^. In particular, since IGS are genetic predictions of imaging traits, we expect to see links between IGS and various phenotypes when there are shared genetic effects.

Here, we systematically developed and evaluated IGS for UKB brain and body images. We examined 4,206 brain IDPs from various imaging modalities and independent processing pipelines, including 3,905 traits generated by the UKB brain imaging team^1,2,34^ (referred to as UKB-Oxford data hereafter) and 301 traits processed by the Brain Imaging Genetics Knowledge Portal (BIG-KP)^33,35,37^. These imaging biomarkers spanned major brain MRI categories, including structural MRI (sMRI) such as regional brain volumes^45^, cortical thicknesses, and surface areas^1^; diffusion MRI (dMRI) such as diffusion tensor imaging (DTI) parameters^46,47^; resting-state functional MRI (rfMRI) such as independent component analysis (ICA)^1,2,48,49^ or parcellation-based^50^ functional connectivity and activity/amplitude^51^ traits; task-based functional MRI (tfMRI) such as activation z-statistics^1^; and susceptibility weighted brain MRI, incorporating regional median T2*^5^. Moreover, we analyzed 169 body IDPs, including 82 from cardiovascular magnetic resonance (CMR)^3,29^, 41 from abdominal MRI^52–54^ (covering organs such as liver, kidney, and lung), and 46 from optical coherence tomography (OCT) imaging^40^. The **Methods** section provides more detailed information of these 4,375 IDPs (4,206 brain + 169 body) (**Table S1**). **Figure 1A** presents an overview of the study design.

**Fig. 1.**
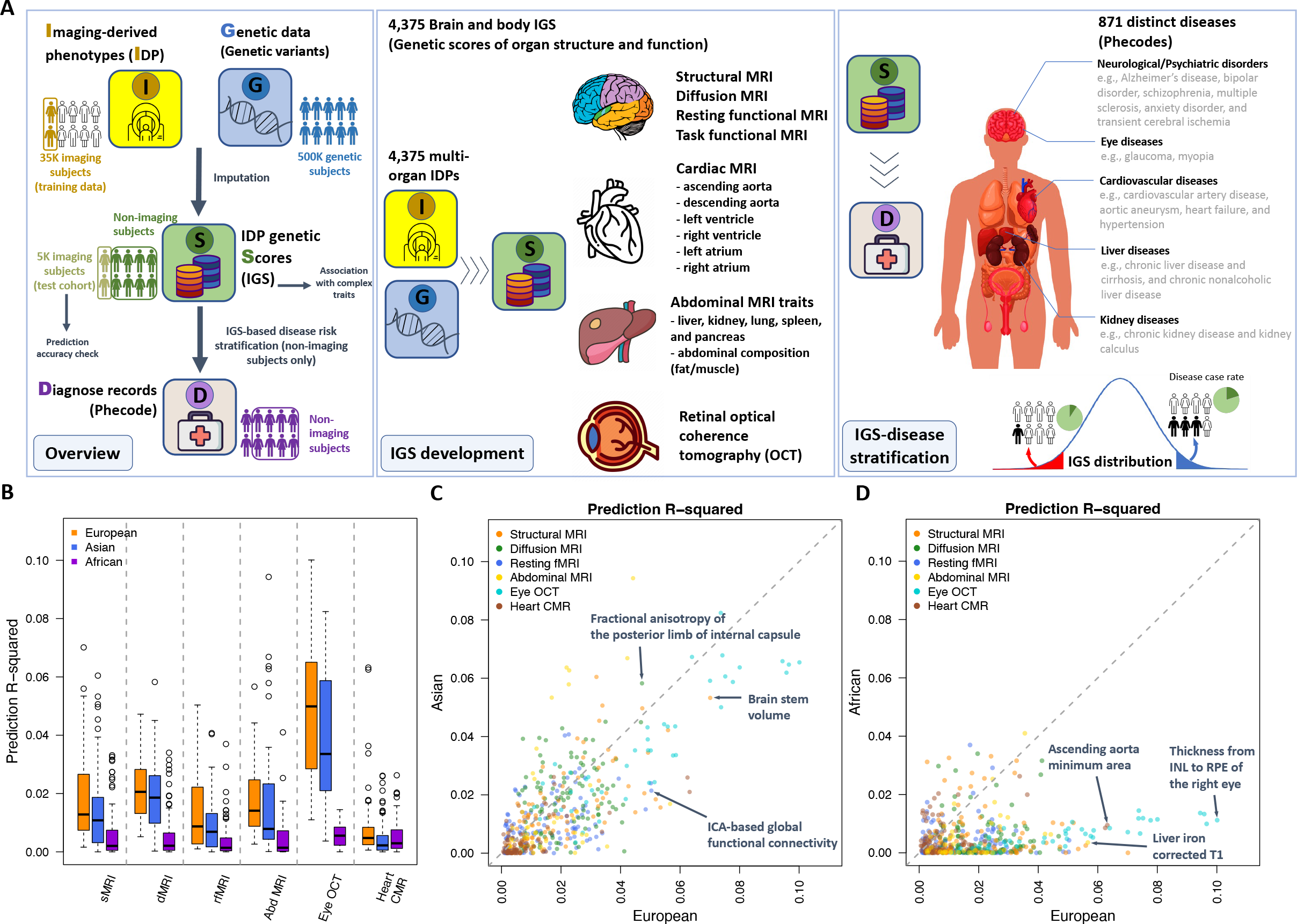
Study overview and prediction analysis**. (A)** An overview of the study design. **(B)** The incremental prediction *R^2^* of brain and body IGS in the hold-out UK Biobank (UKB) testing sets of European, Asian, and African ancestries. The structural MRI (sMRI) modality consisted of 101 BIG-KP regional brain volumes, the diffusion MRI (dMRI) modality consisted of 110 BIG-KP DTI parameters, and the resting functional MRI (rfMRI) modality consisted of 90 BIG-KP parcellation-based traits and 82 UKB-Oxford ICA-based rfMRI traits. The abdominal MRI (Abd MRI) modality consisted of 41 abdominal MRI traits, the eye OCT modality consisted of 46 eye traits, and the heart CMR modality consisted of 82 UKB heart CMR traits. The results of other IGS can be found in **Figure S1**. We display the IGS that significantly predicted the corresponding IDPs in the European dataset after controlling the FDR rate at a 5% level. **(C)** The incremental prediction *R^2^* of these IGS in the European and Asian testing datasets. **(D)** The incremental prediction *R^2^* of these IGS in the European and African testing datasets. In **(C)-(D)**, the IGS with the highest prediction *R^2^* in the European testing set in each modality was marked by an arrow and text. ICA stands for independent component analysis, INL stands for inner nuclear layer, and RPE stands for retinal pigment epithelium.

## RESULTS

### Genetic scores for 4,206 brain IDPs and 169 body IDPs

To develop the genetic scores for IDPs, we used data from UKB individuals of British ancestry with both imaging and genotyping array for training (average *n* = 34,293 for MRI and 54,761 for OCT). An independent hold-out dataset containing participants of non-British European ancestry served as a testing dataset to evaluate the predictive performance of the IGS (average *n* = 5,116 for MRI and 4,801 for OCT). We generated GWAS summary statistics for each IDP, which were then used as input to derive IGS for all UKB participants without imaging data. We used PRS-CS^55^ to construct the IGS with genotyping data, and 461,488 genetic variants were included in the prediction model after standard genetic data quality controls (**Methods**).

We found that 68.14% (2,866/4,206) of brain IDPs and 95.27% (161/169) of body IDPs were significantly predicted in the UKB European testing dataset (Benjamini-Hochberg false discovery rate [FDR] < 5%; **Table S2**). Among the IDPs showing significant prediction, the average *R^2^* was 1.40% (range = [0.10%, 7.62%]) for brain MRI, 0.85% (range = [0.06%, 6.32%]) for heart CMR, 1.80% (range = [0.26%, 5.67%]) for abdominal MRI, and 4.99% (range = [1.10%, 10.01%]) for eye OCT. In the brain, we observed genetically predictable IDPs in both UKB-Oxford and BIG-KP across all brain MRI modalities, and IDPs from the same imaging modality exhibited similar predictive accuracy ranges in the two separate processing pipelines (**Fig. S1A**).

Among BIG-KP sMRI regional brain volumes, the highest prediction accuracy was observed for brain stem volume (*R^2^* = 7.01%). Similarly, the UKB-Oxford sMRI IDP with the highest prediction accuracy (*R^2^* = 7.62%) was the volume of pons by subcortical volumetric sub-segmentation of the brain stem. Among BIG-KP dMRI DTI parameters, the highest prediction accuracy was the mean fractional anisotropy (FA) of the posterior limb of internal capsule (*R^2^* = 4.72%). For rfMRI IDPs, the highest prediction accuracy was observed on one ICA-based global functional connectivity trait that captured the central executive, salience, and default mode networks (*R^2^* = 5.04%). In addition, the IDP with the highest prediction accuracy was the ascending aorta minimum area for heart CMR (*R^2^* = 6.32%) and the liver iron corrected T1 for abdominal MRI (*R^2^* = 7.38%). For eye OCT, the thickness from the inner nuclear layer (INL) to the retinal pigment epithelium of the right eye had the highest prediction accuracy (*R^2^* = 10.01%).

UKB has smaller numbers of non-European participants with imaging data. In an exploratory analysis, we found that many developed IGS exhibited predictive utility for individuals of Asian and African ancestries (average *n* = 460 and 252, respectively), although the prediction accuracy was generally smaller in these cohorts (**Figs. 1B-1D**, **S1B- S1D**, and **Tables S3-S4**). To ensure the robustness of our prediction models, we also conducted the same analyses on all 4,375 IDPs using DBSLMM^56^ and evaluated the consistency of prediction performance (**Figs. S2A-S2B**). Moreover, we performed prediction using imputed genetic data, which required far greater computational resources. We observed similar patterns with improved prediction accuracy (**Figs. S2C- S2D**). In summary, our results provide converging evidence that multi-organ IGS can be reliably developed using different prediction methods and genetic data types, and these IGS can be potentially applied to diverse populations.

### IGS can stratify participants with a higher risk of brain and body diseases

Stratifying participants into different disease risk groups and identifying high-risk individuals is crucial in clinical research and has potential for clinical translation^57–59^. Various factors and biomarkers have been examined for risk stratification, such as disease polygenic risk scores (dPRS)^60–63^, lifestyle factors^64,65^, and imaging traits^66,67^. Here we evaluated the performance of IGS in disease risk stratification for 871 phecode-based^68,69^ diseases (**Table S5**). Considering the comparable prediction accuracy and association results (**Supplementary Note, Figs. S3-S7,** and **Tables S6-S9**) between BIG-KP and UKB- Oxford brain IGS, we focused on a selected set of 383 multi-modality brain IGS together with 41 abdominal IGS, 82 heart IGS, and 46 eye IGS in the following stratification analysis. For both brain and body IGS, we randomly divided 318,781 unrelated UKB discovery (non-imaging) participants into three groups (after removing the effects of covariates): the lowest 10%, the highest 10%, and the middle 80%. We then compared the difference in disease case rate of these three IGS-stratified groups using a chi-squared test^70^ (**Methods**).

We conducted a stratification analysis of 69 brain disorders using 383 brain IGS, which consisted of 101 sMRI IGS (BIG-KP regional brain volumes^33^), 110 dMRI IGS (BIG-KP DTI parameters^35^), and 172 rfMRI IGS (82 ICA-based^2^ and 90 parcellation-based^37^ traits). After controlling FDR at 5% (69 × 383 tests), on UKB discovery cohort, we found that 141 brain IGS stratified 25 disorders into groups with significantly different disease case rates, resulting in a total of 294 IGS-disease pairs, and 247 of them were replicated on the UKB replication cohort (*P* range = [5.31 × 10^-45^, 5.49 × 10^-4^]) (**Figs. 2A-2C** and **S8,** and **Table S10**, **Methods**). Among them, we found that rfMRI IGS accounted for almost two-thirds of the identified stratifications with brain disorders (167/247, 67.61%). The majority (150/167, 89.82%) of the rfMRI IGS-brain disorder stratifications involved delirium, dementia, and Alzheimer’s disease (IGS tail case ratio range = [1.16, 2.46], *P* range = [5.31 × 10^-45^, 5.43 × 10^-4^]) (**Supplementary Note**), and the rest mainly related to multiple sclerosis and cerebrovascular diseases (IGS tail case ratio range = [1.06, 1.92], *P* range = [2.34 × 10^-7^, 5.06 × 10^-4^]). Besides, both the ICA-based and parcellation-based rfMRI IGS displayed a consistent pattern in stratifying delirium, dementia, and Alzheimer’s disease, with the ICA-based traits exhibiting higher power. For example, 14 parcellation-based rfMRI IGS had 38 stratification pairs with delirium, dementia, and Alzheimer’s disease, while 39 ICA- based rfMRI IGS had 119 such pairs, representing 67.86% (38/56) and 90.84% (119/131) of all parcellation-based rfMRI and ICA-based stratification results, respectively (**Table S10**).

**Fig. 2.**
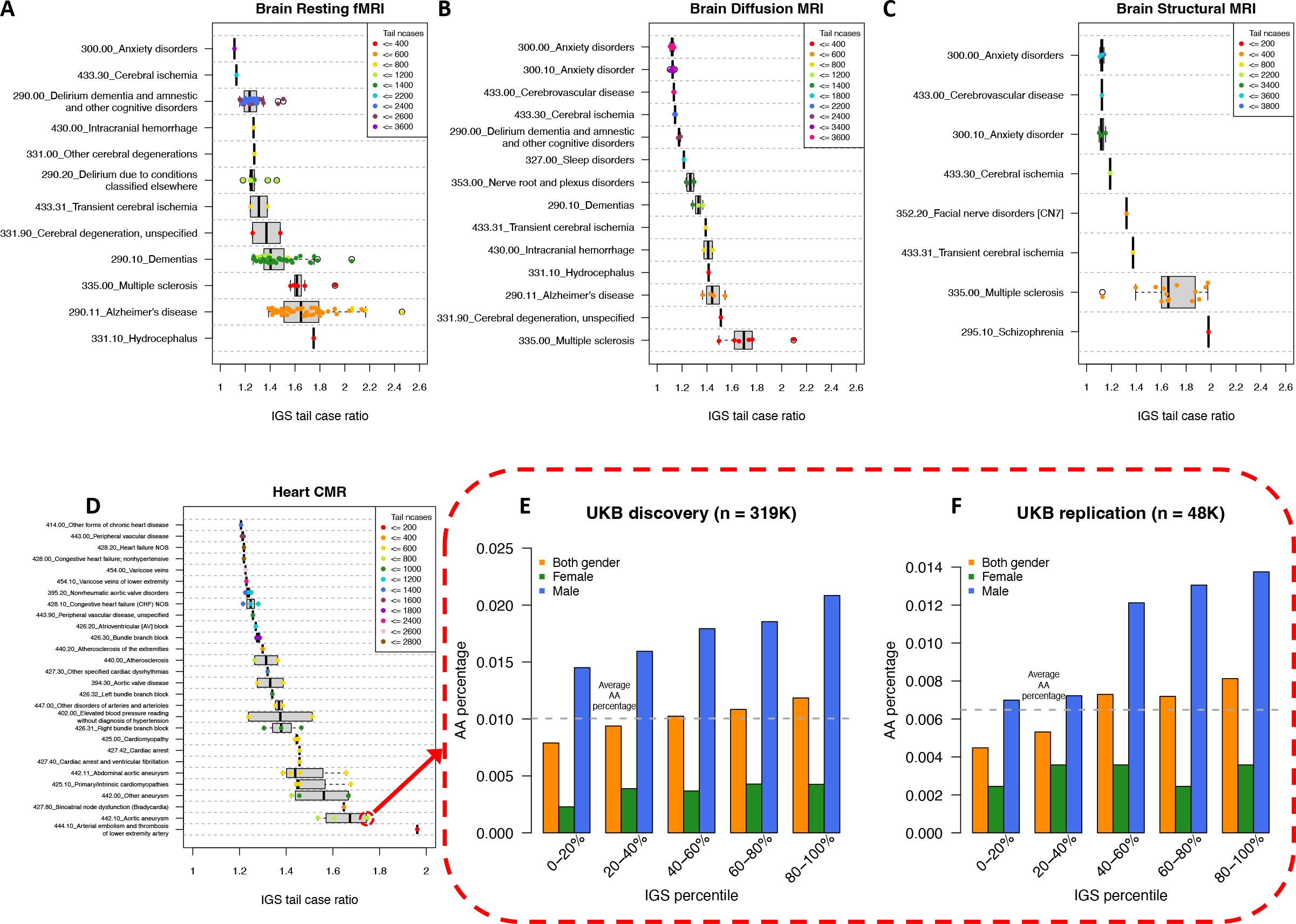
IGS stratification of brain and heart disorders. **(A)-(D)** The IGS-disease pairs after controlling the FDR rate at a 5% level on UKB discovery cohort, replicated on UKB replication cohort, and with IGS tail case ratio greater than 1.1 (brain) or 1.2 (heart) are displayed. Each point represents an IGS-disease pair. The *y*-axis shows the disease name and the associated phecode (for example, the phecode of Alzheimer’s Disease is 290.11). The *x*-axis shows the disease case ratio between the two IGS tails. We show the results for UKB discovery cohort. **(E)-(F)** For the pair between aortic aneurysm (AA) and AAo_max_area (ascending aorta maximum area), we show the disease percentage (*y*-axis) in quantile-based groups defined by AAo_max_area IGS (*x*- axis) in the UKB discovery and UKB replication cohorts. The horizontal grey dashed line represents the disease percentage for the entire cohort.

The IGS derived from dMRI and sMRI also stratified multiple brain disorders. Out of the 110 dMRI IGS, 33 significantly stratified 14 disorders (IGS tail case ratio range = [1.04, 2.10], *P* range = [1.65 × 10^-8^, 5.12 × 10^-4^]), corresponding to 54 IGS-disease pairs, including multiple sclerosis, dementia, Alzheimer’s disease, and cerebrovascular diseases such as intracranial hemorrhage, cerebral ischemia, and occlusion of cerebral arteries (**Fig. 2B**). Among the 101 sMRI IGS, 14 significantly stratified 9 disorders (IGS tail case ratio range = [1.09, 1.98], *P* range = [3.50 × 10^-11^, 5.49 × 10^-4^]), with a total of 26 IGS-disease pairs (**Fig. 2C**). We found that both dMRI and sMRI IGS showed strong stratifications with multiple sclerosis, a heritable common neurodegenerative disease and a leading cause of nontraumatic disability among young adults^71–73^. For example, the IGS of the cingulum axial diffusivity was found to have significantly more MS patients in its lower 10% tail than its upper 10% tail (IGS tail case ratio = 1.76, *P* = 3.45 × 10^-8^), consistent with results of previous association analysis using dMRI data^74^. Participants with the smallest 10% IGS of the left and right thalamus proper volumes contained 1.73 and 1.96-fold (*P* < 5.37 × 10^-5^) more MS cases than those with the largest 10% IGS, respectively. Similarly, IGS of the left and right putamen and the accumbens area volumes all stratified MS patients such that the lower 10% IGS tail had a significantly higher case number than the upper 10% IGS tail (IGS tail case ratio range = [1.55, 1.85], *P* range = [2.09 × 10^-10^, 4.11 × 10^-4^]). These findings are in line with previous studies identifying decreased brain volume in the thalamus^75,76^, putamen^77^, and accumbens^78^ in multiple sclerosis patients, highlighting the potential of these brain IGS in identifying individuals at high risk for multiple sclerosis without actual imaging data.

Body IGS also stratified diseases in their corresponding organs and systems. Using the 41 abdominal IGS, we stratified 104 digestive system diseases and 100 genitourinary system diseases (204 × 41 tests) and identified 280 significant IGS-disease pairs after controlling FDR at 5% level, where 229 of them were replicated on the UKB replication cohort, associated with 37 abdominal IGS and 51 diseases (*P* range = [4.80 × 10^-250^, 1.66 × 10^-3^]) (**Fig. S9A**). For example, we found that chronic renal failure patients were significantly stratified by the IGS of kidney volume and kidney parenchyma volume (IGS tail case ratio range = [1.25, 1.37], *P* range = [2.05 × 10^-18^, 6.42 × 10^-10^]). The lower 10% IGS tail contained more patients, and indeed the kidney volume and kidney parenchyma volume have been found associated with kidney function (such as positive correlation with as glomerular filtration rate and negative correlation with creatinine)^79,80^. In addition, patients with chronic liver disease and cirrhosis were significantly stratified by the IGS of several abdominal MRI traits, including liver proton density fat fraction and total abdominal fat (IGS tail case ratio range = [1.22, 1.99], *P* range = [4.88 × 10^-40^, 1.55 × 10^-3^]), consistent with previous studies on the association between liver disease risk and liver/abdominal fat^81–83^. Furthermore, we examined the stratification of 90 circulatory system diseases using 82 heart IGS (90 × 82 tests), discovered 202 IGS-disease pairs after controlling FDR at 5% level, and replicated 180 pairs between 54 heart IGS and 40 diseases (*P* range = [2.40 × 10^-34^, 1.37 × 10^-3^]) (**Fig. 2D**). The strongest stratifications were found between aortic aneurysms and the IGS of ascending/descending aorta maximum area, and ascending/descending aorta minimum area (IGS tail case ratio range = [1.32, 1.75], *P* range = [5.71 × 10^-14^, 6.75 × 10^-4^]), which is the area of the region where aortic aneurysm occurs^84^. We found that aortic aneurysm was monotonely stratified across all values of the IGS of the ascending aorta maximum area such that larger IGS corresponded to higher risk (**Figs. 2E-2F**). MRI-measured aortic dimensions are frequently used for diagnosis of aortic aneurysms^85^, and large aortic diameters are considered abnormal and even diagnostic of aortic aneurysms^86^. Similarly, we stratified 38 eye and adnexa-related disorders using 46 eye IGS (38 × 46 tests) and replicated 67 of the 107 IGS-disease pairs with significant stratifications, corresponding to 27 eye IGS and 13 eye disorders (*P* range = [5.65 × 10^-49^, 2.25 × 10^-3^]) (**Fig. S9B**). For example, we found significantly more glaucoma cases presented in the 10% upper tail of the IGS of the raw vertical cup-to-disc ratio (VCDR) of the left eye and the VCDR regressed on disc diameter^87^ (IGS tail case ratio range = [1.51, 1.98], *P* range = [3.41 × 10^-22^, 3.77 × 10^-9^]). The large value of observed VCDR has been used clinically as a robust indicator for glaucoma^88^.

We also evaluated the performance of IGS in cross-organ applications. First, we stratified 69 brain disorders by the 41 abdominal IGS, 82 heart IGS, and 46 eye IGS. We identified and replicated 19 pairs between 19 IGS (6 heart IGS and 13 abdominal IGS) and 9 brain disorders (*P* range = [4.16 × 10^-13^, 9.97 × 10^-5^]) (**Fig. S9C** and **Table S11**). For example, the IGS of the right ventricular ejection fraction had 1.62-fold more multiple sclerosis patients in its lower 10% tail than its upper 10% tail (*P* = 7.03 × 10^-6^). This result matched previous findings that multiple sclerosis patients had decreased ventricular ejection fractions^89,90^. Second, we stratified 90 circulatory system diseases by the 383 brain IGS, 41 abdominal IGS, and 46 eye IGS. After controlling FDR at 5% level and replicating on UKB replication cohort, we found that 27 circulatory system diseases were significantly stratified by 13 abdominal IGS, 79 brain IGS, and 3 eye IGS (*P* range = [1.04 × 10^-10^, 2.78 × 10^-4^]) (**Fig. S9D**). Among these, peripheral vascular disease was widely stratified by all modalities of brain IGS (IGS tail case ratio range = [1.10, 1.36], *P* range = [2.25 × 10^-8^, 2.71 × 10^-4^]), with the majority of signals in dMRI IGS (56.36%, 31/55). For example, the FA IGS of the body of corpus callosum and anterior corona radiata both had more cases in their upper 10% tail, and the mean diffusivity IGS of these two had fewer cases in their upper 10% tail, consistent with prior findings on the associations between dMRI traits and peripheral vascular diseases^91^. Additionally, the IGS of the thickness of the inner nuclear of both eyes significantly stratified hypertension patients, consistent with the reported associations between higher blood pressure and thicker INL^92^, as well as our association results between eye IGS and quantitative measures of blood pressure (**Supplementary Note**).

### IGS broadly contribute to disease stratification in addition to disease genetic risk

Following disease stratification by IGS, we measured the predictive power of IGS for disease by area under the curve (AUC), as AUC was widely used to measure power of PRS^93,94^ and allowed us to better measure the clinical utility of IGS. For each IGS-disease pair that showed significant stratification (**Figs. 2A-2D** and **S9**, and **Tables S10-S11**), we evaluated the AUC gain by including the IGS for disease prediction, compared to a baseline model. Since dPRS is a frequently used genetic risk measure, we also analyzed the AUC gain from the additional inclusion of IGS when FinnGen^95^-derived dPRS was already incorporated (**Methods**).

As previously shown, brain and body IGS demonstrated marginal stratification capability for diseases in both same-organ and cross-organ applications (**Figs. 2A-2D** and **S10**). To measure the AUC gain from IGS, for each of the 110 diseases significantly stratified by IGS and for each IGS that significantly stratified the disease, we constructed two logistic regression models to measure the AUC gain: a baseline model that predicts disease status using basic covariates only, and an IGS model that additionally uses an IGS. The increment in the AUC of the IGS model relative to the baseline model is defined as the AUC gain of IGS. Out of the 919 pairs IGS-disease pairs showing replicated stratification patterns, 916 pairs had positive AUC gain for disease prediction from the IGS. Comparing the baseline model to the IGS model using DeLong test^96^, after controlling for FDR at level 5%, 80.52% (740/919) pairs had significant AUC gain with the median being 0.0208 (range = [7.25 × 10^-5^, 2.11 × 10^-1^], **Figs. 3A-3C** and **S10**, and **Tables S12-S13**). For example, in **Figure 3A**, the largest AUC gain from brain IGS for brain disorder prediction was achieved by Net25_Node9 IGS for Alzheimer’s disease (AUC gain = 0.0858, *P* = 8.04 × 10^-23^), whose disease stratification patterns were visualized in **Figure S8**. All dMRI IGS that stratified Alzheimer’s disease in **Figure 2B** also had significant AUC gain (**Fig. 3B**). The IGS of the ascending aorta maximum area predicted aortic aneurysm with AUC gain 0.0462 (*P* = 5.93 × 10^-9^), the largest among heart IGS that significantly stratified circulatory system diseases (**Fig. 3C**). Overall, the AUC gains of IGS suggest that the majority of disease-stratifying IGS not only stratify subsets of subjects at high risk but also contribute to disease prediction at the cohort level.

**Fig. 3.**
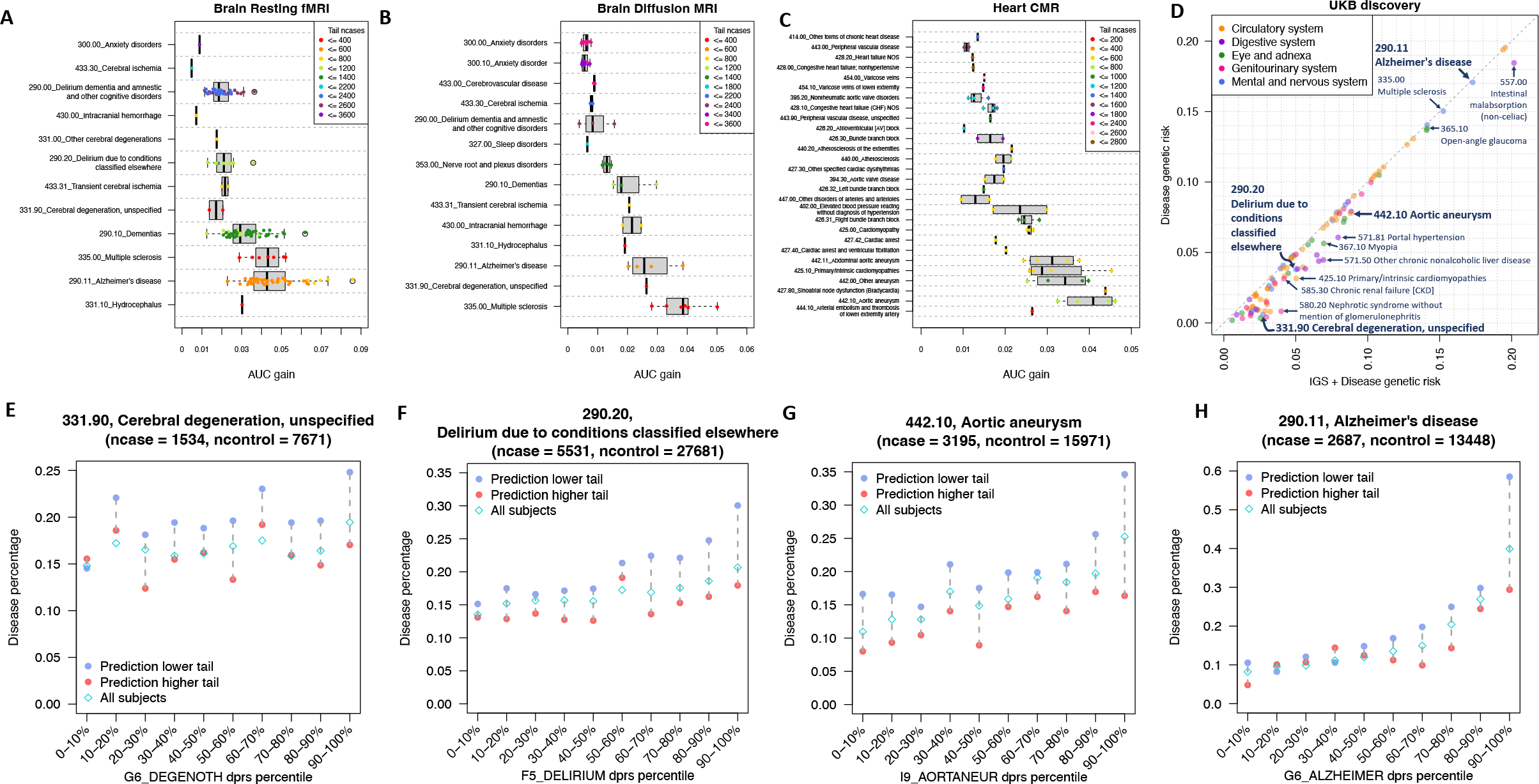
AUC of IGS for disease prediction. **(A)-(C)** AUC gain of disease prediction on UKB discovery cohort by including the stratifying IGS for IGS-disease pairs that were significant on UKB discovery cohort after controlling the FDR rate at a 5% level, replicated on UKB replication cohort, and with IGS tail case ratio greater than 1.1 (brain) or 1.2 (heart). Each point represents an IGS-disease pair. The *y*-axis shows the disease name and the associated phecode. The *x*-axis shows the AUC gain from the IGS model compared with the baseline model. **(D)** AUC gain of disease prediction on UKB discovery cohort by including the most stratifying IGS or all stratifying IGS in the IGS-dPRS model, compared with the baseline-dPRS model. The diseases with IGS stratification patterns visualized in **Figures 3E-3H and S11** was marked by an arrow and text. **(E)-(H)** For four select diseases, for each disease polygenic risk score (dPRS) quantile group (*x*-axis), we show the disease percentage (*y*-axis) for all participants (“All subjects”), participants in the low 10% tail of the prediction of the IGS-dPRS model of the disease (“Prediction lower tail”), and participants in the high 10% tail of the prediction of the IGS-dPRS model (“Prediction higher tail”). We show the results for UKB discovery cohort.

We further assessed the contribution of IGS to disease prediction and stratification beyond that provided by dPRS. Briefly, we constructed dPRS using FinnGen GWAS summary statistics of phecode-based diseases. We manually mapped 105 of the 110 UKB diseases that had IGS stratification signals to 78 FinnGen disease endpoints (case number range = [216, 111,581], median = 7,576) following the FinnGen-phecode mapping in Sun et al^97^ and disease description in FinnGen R9^95^ (**Table S14**). After Bonferroni correction at 5% level, 98 out of the 105 diseases gained significantly in AUC from inclusion of IGS on top of dPRS (AUC gain range = [5.85 × 10^-5^, 3.16 × 10^-2^], AUC gain median = 4.26 × 10^-3^, *P* range = [7.24 × 10^-35^, 2.27 × 10^-3^]) (**Fig. 3D** and **Table S15**). The largest AUC gain (0.0316) was achieved for nephrotic syndrome (without mention of glomerulonephritis, phecode 580.20), a kidney disorder associated with excessive protein in the urine. Examining the AUC from dPRS alone and the additional AUC gain from IGS in **Figure 3D**, diseases can be categorized into three types: (1) those with uninformative dPRS and large relative AUC gains from IGS, (2) those with moderate dPRS and moderate AUC gains from IGS, and (3) those with informative dPRS and minimal AUC gains from IGS. This suggests that IGS can help disease stratification, even if it does not always substantially boost the AUC. It makes sense because AUC primarily reflects overall predictive performance across the entire cohort and might not capture variations within specific subgroups (e.g., subjects with high/low risk). Multiple diseases from each category were discussed in **Supplementary Note**, and below we highlight a few examples spanning all categories.

For diseases with “uninformative dPRS,” where subjects may have similar disease risks across all dPRS percentile groups, IGS can still effectively stratify these subjects into high and low risk within each dPRS group (**Fig. 3E**). Typically, these diseases lack large GWAS data resources, and incorporating IGS could substantially improve the AUC. For example, the dPRS of nephrotic syndrome had an AUC gain of 0.0083 and was generated using FinnGen GWAS (FinnGen ID: N14_NEPHROTICSYND), which had a limited sample size of 853 patients but was already large among the existing GWAS of this disease^98,99^. Cerebral degeneration (phecode 331.90) had a dPRS AUC gain of 0.0084 (G6_DEGENOTH) and gained additional AUC of 0.0188 by including three brain IGS. As the predictive performance of dPRS improved, resulting in “moderate dPRS,” the stratification ability of the IGS-dPRS model remained consistent on top of dPRS across almost all dPRS groups (**Figs. 3F-3G**). IGS AUC gain remained nontrivial for some diseases, such as delirium (phecode 290.20, AUC gain 0.0122) and aortic aneurysm (phecode 442.10, AUC gain 0.0101). Furthermore, Alzheimer’s disease and multiple sclerosis had “informative dPRS” such that the dPRS AUC gain was 0.1707 and 0.1503, respectively, and thus the further AUC gain from IGS was significant yet small, both being 0.0022. Nevertheless, individuals with high Alzheimer’s disease dPRS, especially the top 10% individuals with highest dPRS, were stratified into subgroups of distinct disease risk by IGS (**Fig. 3H**). Similar stratification trends from the IGS-dPRS model were observed across multiple sclerosis and various other diseases (**Fig. S11**).

In summary, we conducted a comprehensive AUC analysis of IGS prediction of diseases. Most disease-stratifying IGS showed significant AUC contribution, and various diseases, such as nephrotic syndrome, cerebral degeneration, delirium, and aortic aneurysms, had nontrivial AUC gain from IGS in addition to dPRS. Besides, even for diseases with informative dPRS and seemingly minimal AUC gain from IGS, IGS still stratified individuals with high disease genetic risk into subgroups with distinct risk. We will conduct detailed analyses of IGS stratification for Alzheimer’s disease and multiple sclerosis in the next section to provide more insights into the IGS contribution to disease stratification in scenarios with minimal AUC gains. It is important to note that these cases may represent the lower bound of IGS utility, as their contribution would likely be greater if the AUC gains were more substantial.

### IGS stratification of Alzheimer’s disease and multiple sclerosis: in-depth analysis and generalizability

As presented earlier, Alzheimer’s disease and multiple sclerosis are the two neurodegenerative disorders that were most frequently stratified by brain IGS on UKB discovery cohort, with 55 and 27 stratifying brain IGS, respectively. In this section, we provide an in-depth analysis of the stratification patterns of these two diseases using all stratifying brain IGS. Furthermore, we constructed IGS for participants from two independent cohorts: the All of Us research program (AOU, *n* = 245,394)^100^ and Alzheimer’s Disease Neuroimaging Initiative study (ADNI, *n* = 1,152)^101^ (**Methods**). We showed that the Alzheimer’s disease stratification patterns observed on UKB can be replicated on AOU and ADNI. Moreover, we considered *APOE* status, and found that IGS can effectively stratify Alzheimer’s disease among participants of same *APOE* haplotypes. *APOE* is known to be the strongest genetic factor risk for Alzheimer’s disease and its homozygote is considered a form of Alzheimer’s disease^102^. Similar to Alzheimer’s disease, multiple sclerosis stratification patterns on UKB were replicated on AOU as well.

We have observed the marginal stratification of Alzheimer’s disease on UKB participants by many brain IGS (**Figs. 2A-2B**). To stratify Alzheimer’s disease using all the stratifying IGS, we constructed an IGS burden score by counting the number of IGS high-risk tails a subject was in, across the 55 IGS that were marginally stratifying (**Methods**). As expected, Alzheimer’s disease risk rose as IGS burden score increased (**Fig. S12**). Two additional risk factors, age and dPRS, were then evaluated. Ageing is known to be closely related to Alzheimer’s disease progression^103,104^. Upon grouping participants by age, participants of similar age were further stratified into “lower IGS burden score” or “higher IGS burden score” groups if their IGS burden scores fell within the lower or upper 20% extremes for their respective stratum. We found a consistent rise in the percentage of disease cases with increasing age. Within each age group, particularly beginning from around age 60 on the UKB cohorts and from around age 65 on the AOU and ADNI cohorts, participants with higher IGS burden score always had much higher disease percentage than those with lower IGS burden score (**Figs. 4A-4D** and **Table S16**).

**Fig. 4.**
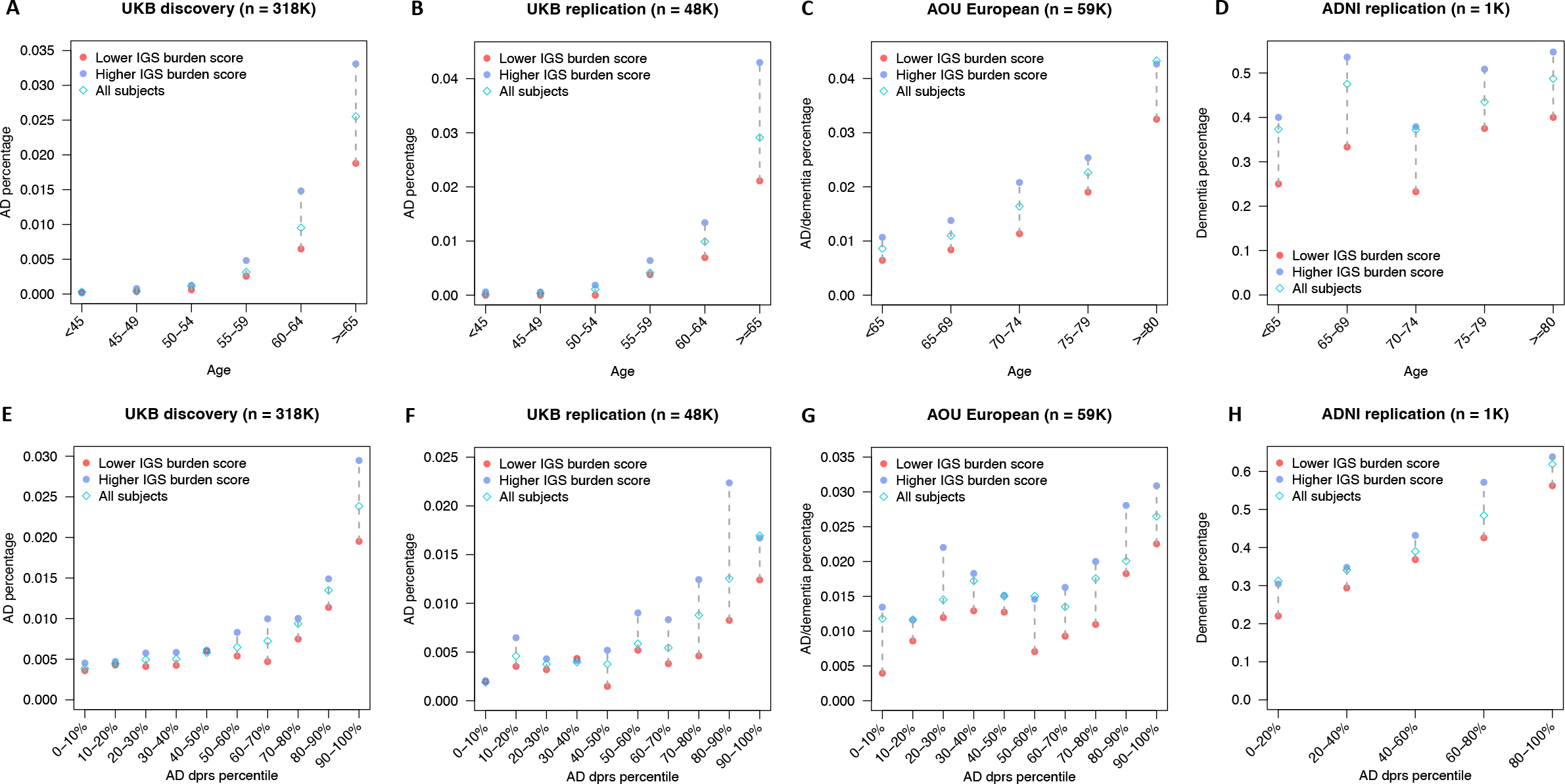
IGS stratification of Alzheimer’s disease across age and disease polygenic risk score groups. **(A)-(D)** For each age group (*x*-axis), we show the Alzheimer’s disease (AD) or dementia percentage (*y*-axis) for all participants (“All subjects”), participants in the low 20% tail of IGS burden score (“Lower IGS burden score”), and participants in the high 20% tail of IGS burden score (“Higher IGS burden score”). **(E)-(H)** For each dPRS quantile group (*x*-axis), we show the AD or dementia percentage (*y*-axis) for all participants (“All subjects”), participants in the low 20% tail of IGS burden scores (“Lower IGS burden score”), and participants in the high 20% tail of IGS burden scores (“Higher IGS burden score”). We show the results for UKB discovery (left panels), UKB replication (middle-left panels), AOU European (middle-right panels), and ADNI (right panels) cohorts.

For Alzheimer’s disease dPRS, we first verified that the FinnGen-derived dPRS effectively stratified patients in UKB, AOU, and ADNI cohorts (**Fig. S13**). We found that the IGS burden score further stratified participants from the same dPRS percentile group. For example, in **Figure 4E**, among the 31,797 participants with the top 10% dPRS, 2.38% had developed Alzheimer’s disease. The subgroup with higher IGS burden scores had an disease percentage of 3.09% and the one with lower IGS burden scores had a disease percentage of 1.85%. That is, inside this dPRS group, the higher IGS burden score subgroup contained 1.67-fold of cases, compared with the lower IGS burden score subgroup (*P* = 2.58 × 10^-6^). Similar patterns were observed in UKB replication and AOU cohorts (**Figs. 4F-4G**). In ADNI, we also found that within each dPRS percentile group, the subgroup of higher IGS burden score always had a higher dementia percentage than the lower IGS burden score subgroup (**Fig. 4H**). These results demonstrated the contribution of IGS stratification on top of dPRS and the generalizability of UKB-derived IGS on external genetic cohorts.

We then examined *APOE* and found the IGS stratification power was not dominated by *APOE* status. In the UKB discovery cohort, we focused on subjects of age at least 55, and belong to one of the four common APOE haplotypes (*n* = 202,304): ε2 carriers, ε3 homozygotes, ε4 heterozygotes, and ε4 homozygotes (**Methods**). ε3 homozygotes were the most common *APOE* haplotype, ε2 carriers had lower disease risk, ε4 heterozygotes had higher risk, and ε4 homozygotes were recently considered a genetic form of Alzheimer’s disease^102^. ε4 heterozygotes and ε4 homozygotes were also combined into ε4 carriers. In **Figure 5A**, we found that IGS burden score stratified disease risk among participants of same *APOE* haplotypes, and IGS stratification power remained when both disease dPRS and *APOE* status were considered (**Figs. 5B-5F**). These results demonstrated that IGS stratification of Alzheimer’s disease was not simply driven by *APOE*, suggesting that IGS could be useful for risk stratification when combined with other genetic risk factors.

**Fig. 5.**
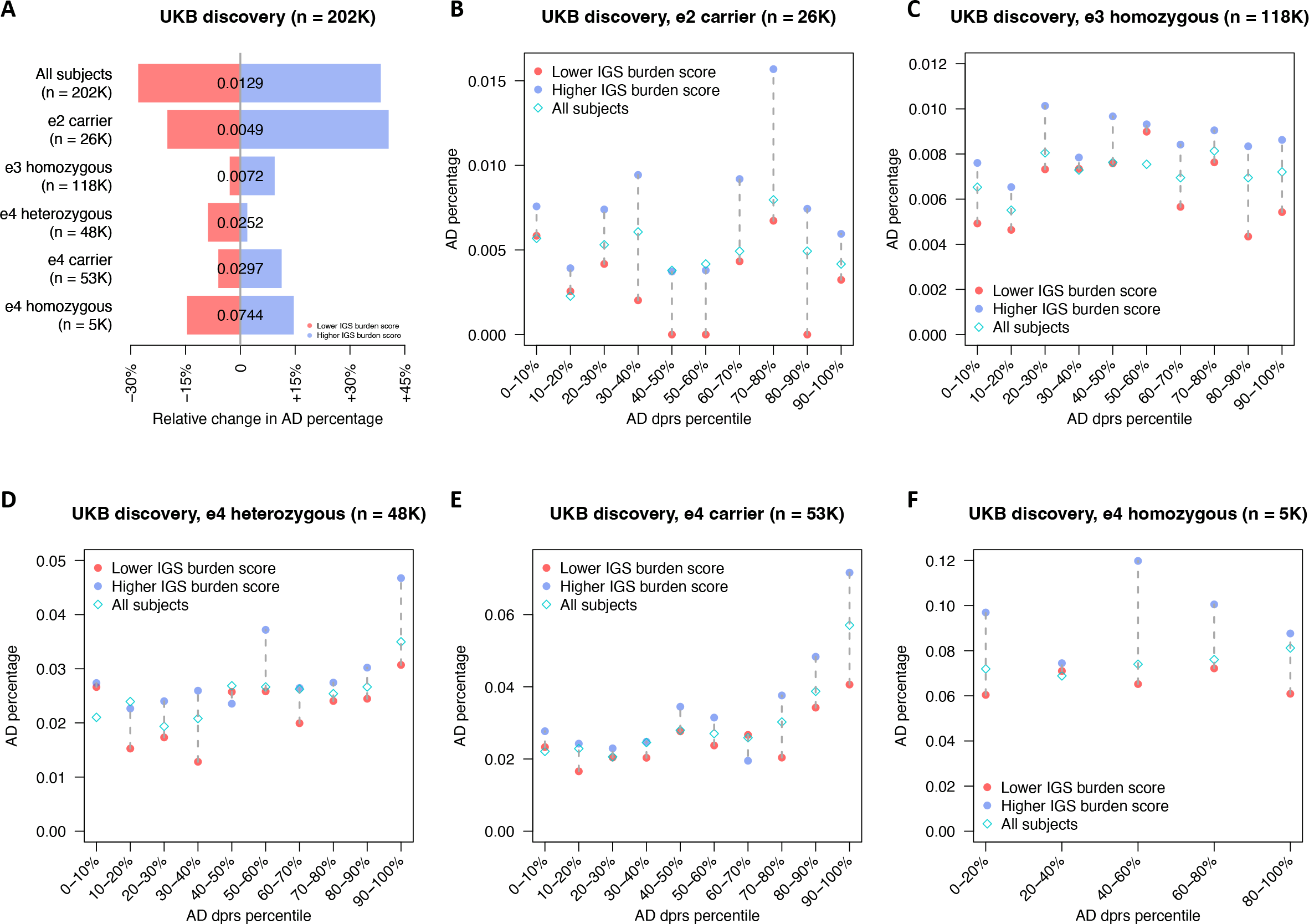
IGS stratification of Alzheimer’s disease on the basis of *APOE* status and disease polygenic risk score. **(A)** For each APOE group (*y*-axis), we show the relative change in Alzheimer’s Disease (AD) percentage (*x*-axis) in the low 20% tail of IGS burden score (“Lower IGS burden score”) and in the high 20% tail of IGS burden score (“Higher IGS burden score”) compared to the overall AD percentage of the APOE group (decimals at *x*=0). **(B)-(F)** For five *APOE* groups, for each dPRS quantile group (*x*-axis), we show the AD percentage (*y*-axis) for all participants (“All subjects”), participants in the low 20% tail of IGS burden score (“Lower IGS burden score”), and participants in the high 20% tail of IGS burden score (“Higher IGS burden score”). We show the results for the UKB discovery cohort.

Brain IGS and burden scores also stratified multiple sclerosis on both UKB and AOU cohorts. For example, the IGS of the left putamen volume had 1.85-fold of multiple sclerosis patients in its lower 10% tail than its upper 10% tail (**Fig. 2C** and **Table S10**). Decreased putamen volume was observed among multiple sclerosis patients in previous studies^77^. Further examining participants across all IGS percentile groups, we found that both UKB discovery participants and AOU European participants had roughly monotone decreasing disease percentage as the left putamen volume IGS increased, with the monotone trend more visible among females (**Figs. 6A-6B** and **Table S17**). We then included dPRS as an additional risk factor to stratify multiple sclerosis. Similar to Alzheimer’s disease dPRS, we first verified that multiple sclerosis dPRS were able to stratify multiple sclerosis patients in UKB and AOU cohorts (**Fig. S14**). IGS burden score was then constructed and was able to further stratify participants of similar dPRS into subgroups of different disease percentage (**Methods**). Among both UKB and AOU participants, further stratification of multiple sclerosis by IGS burden score on top of dPRS was observed, and the stratification was weaker on non-European cohorts as expected (**Figs. 6C-6F**). The multiple sclerosis stratification patterns on UKB and AOU using UKB- derived IGS provided evidence that stratification patterns by UKB-derived IGS could generalize well to external cohort, especially when the ancestry of the external cohort is close the IGS development cohort. We also examined stratification of bipolar disorder in the AOU study (**Supplementary Note** and **Table S18**), which represents a scenario where the external cohort has a much larger number of disease cases compared to the UKB study.

**Fig. 6.**
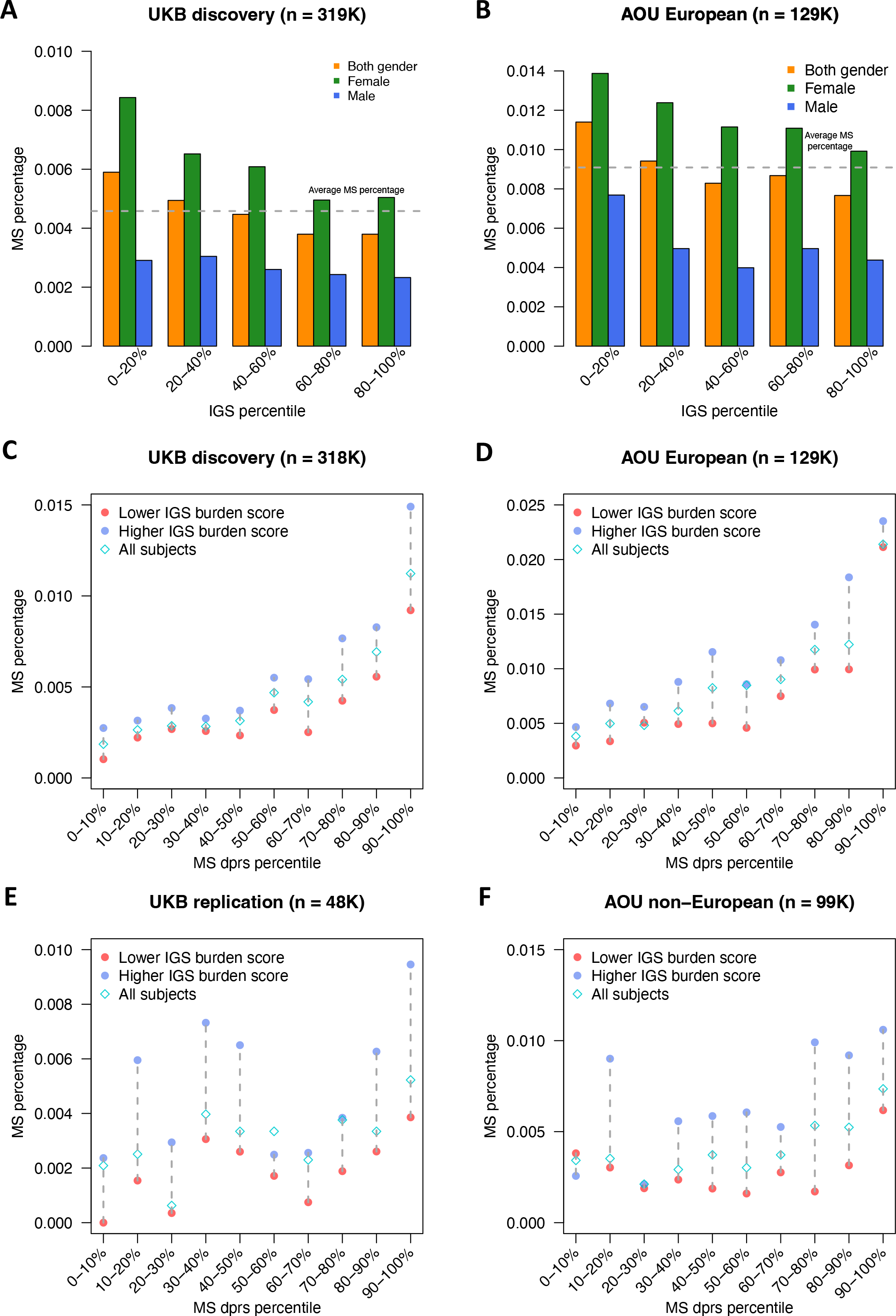
IGS stratification of multiple sclerosis. **(A)-(B)** For the pair between multiple sclerosis (MS) and left.putamen (left putamen volume), we show the disease percentage (*y*-axis) in quantile-based groups defined by left.putamen IGS (*x*-axis) in the UKB discovery and AOU European cohorts. The horizontal grey dashed line represents the disease percentage for the entire cohort. **(C)-(E)** For each dPRS quantile group (*x*-axis), we show the MS percentage (*y*-axis) for all participants (“All subjects”), participants in the low 20% tail of IGS burden scores (“Lower IGS burden score”), and participants in the high 20% tail of IGS burden scores (“Higher IGS burden score”). We show the results for UKB discovery, UKB replication, AOU European, and AOU non-European cohorts.

## DISCUSSION

In this study, we developed 4,375 brain and body IGS and evaluated their performance in disease risk stratification for UKB participants without imaging data and external participants from UKB-independent studies. We confirmed that these IGS can provide biologically relevant information to complex traits and diseases. Brain and body IGS effectively stratified the risk of various diseases not only within their respective organ systems but also in other organs, such as the pair of right ventricular ejection fraction and multiple sclerosis. These results reflect both intra- and inter-organ connections between IGS and clinical outcomes. We also conducted AUC analysis of IGS and found IGS can stratify disease risk on top of disease genetic risk score, regardless of the overall AUC gain. These results highlight differences in AUC measures and stratification capabilities, suggesting the potential of IGS for clinical utility. We conducted a thorough analysis of Alzheimer’s disease, detailing age-related trends, replication of findings in both UKB and independent cohorts, and the distinct contributions of IGS beyond dPRS and *APOE*. Our IGS data resources have been made publicly available, facilitating similar in-depth analyses for numerous brain and somatic diseases. We also provide a second example of this application to multiple sclerosis.

Ageing is well-recognized for its close association with the progression of Alzheimer’s disease. Our findings indicate that IGS burden score consistently stratifies individuals at higher risk of Alzheimer’s disease in those aged over 55 in UKB and over 65 in AOU and ADNI (**Figs. 4A-4D**). The IGS burden score was derived from multiple brain MRI IDPs, many of which have been previously associated with Alzheimer’s disease in studies with actual imaging. Given that IGS was constructed solely from genotyping data, it can be readily applied to participants with genetic data, enabling a quick and efficient assessment of disease risk in older individuals, without undergoing real imaging data collection. We further demonstrated that IGS, when combined with dPRS, offered a multi-layered stratification of disease risk, which had better performance than using only the disease genetic risk score (**Figs. 4E-4H**). Moreover, IGS contributed to Alzheimer’s disease stratification when both *APOE* status and dPRS were considered (**Figs. 5A-5F**), suggesting stratification power of IGS was not purely driven by *APOE*. These results indicate that although the UKB imaging study, with its largely healthy participant base, may not have direct proven applicability for disease risk management, it possesses the capacity to stratify disease risk in broader external genetic cohorts. We anticipate that the IGS developed in our study can be widely applied in the stratification and management of Alzheimer’s disease risk.

When real imaging data are not available, IGS can be used as genetically predicted variables for brain and body structures and functions. In addition to contribute to disease stratification and prediction, established IGS provides an objective pseudo-imaging biomarker associated with a specific organ region, which may serve as informative measure of diseases such as early-stage Alzheimer’s disease^105,106^, early-stage multiple sclerosis^107,108^, and bipolar disorder^109^. Furthermore, IGS may also contribute to ongoing efforts on disease subtyping and integration^110,111^. However, as shown in our prediction analysis, IGS was typically only able to partially reconstruct the imaging phenotypes. It has generally been observed that IGS has demonstrated imperfect performance in most complex traits and diseases, which can be attributed to many factors, including a limited number of training samples, heritability, and weak genetic effects^112–114^. Another challenge in IGS applications lies in ancestry and population differences^115–117^. As the current UKB imaging cohort had the majority of the participants of European ancestry, generating IGS in non-UKB and/or non-European studies may have further reduced performance^113^. In our UKB replication cohort, comprising non-British and non-European participants, we observed that the IGS demonstrated consistent performance similar to that in the discovery cohort (for example, **Figs. 4B, 4F,** and **6E**). More powerful IGS methods that better account for these IGS limitations and cohort differences may result in more informative IGS for potential clinical applications. In addition, we expect to see improvement in IGS prediction accuracy and stratification when more imaging samples and IDPs are released by the UKB and other large imaging genetic studies.

## METHODS

Methods are available in the ***Methods*** section.

## ACKNOWLEDGEMENTS

Research reported in this publication was supported by the National Institute On Aging of the National Institutes of Health under Award Number RF1AG082938. The content is solely the responsibility of the authors and does not necessarily represent the official views of the National Institutes of Health. The study has also been partially supported by funding from the Wharton Dean’s Research Fund, Analytics at Wharton, and Purdue Statistics Department. This research has been conducted using the UK Biobank resource (application number 76139), subject to a data transfer agreement. We would like to thank the individuals who represented themselves in the UK Biobank for their participation and the research teams for their efforts in collecting, processing, and disseminating these datasets. We gratefully acknowledge all the studies and databases that made GWAS summary-level data publicly available. We would like to thank the research computing groups at Purdue University, the University of North Carolina at Chapel Hill, and the Wharton School of the University of Pennsylvania for providing computational resources and support that have contributed to these research results.

## METHODS

### Imaging traits

The imaging data used in our study were obtained from the UKB study, which recruited around 500,000 individuals between the ages of 37 and 73 (229,114 males) between 2006 and 2010^118^ (https://www.ukbiobank.ac.uk/). The ethics approval of the UKB study was obtained from the North West Multicentre Research Ethics Committee (approval number: 11/NW/0382). We used a total of 4,206 brain MRI IDPs, 41 abdominal MRI IDPs, 82 heart CMR IDPs, and 46 eye OCT IDPs. The brain MRI IDPS consisted of 301 BIG-KP^33,35,37^ and 3,905 UKB-Oxford^1,2,34^ traits. The BIG-KP traits were from our previous studies on three brain MRI modalities. First, we obtained 101 regional brain volumes^33^ from sMRI by applying the advanced normalization tools^45^. Second, we generated 110 tract-averaged DTI parameters^35^ from dMRI using the ENIGMA-DTI pipeline^46,47^. Third, for rfMRI, we partitioned the cerebral cortex into 360 brain areas using the Glasser360 parcellation^50^. We obtained 90 functional activity (amplitude) and functional connectivity (edge) traits^37^ for 12 functional networks^119^. The UKB-Oxford had 1,437 IDPs from sMRI (including susceptibility-weighted structural imaging), 675 from dMRI, 1,777 from rfMRI, and 16 from task-based tfMRI. Specifically, the sMRI IDPs consisted of FIRST (Category 1102), FAST (Category 1101), FreeSurfer ASEG (Category 190), FreeSurfer BA exvivo (Category 195), FreeSurfer a2009s (Category 197), FreeSurfer DKT (Category 196), FreeSurfer desikan gw (Category 194), FreeSurfer desikan pial (Category 193), FreeSurfer desikan white (Category 192), FreeSurfer subsegmentation (Category 191), regional T2* (Category 109), and white matter hyperintensity volume (Category 112). The 675 dMRI IDPs included 432 TBSS-processed IDPs from Category 134 and 243 ProbtrackX-processed IDPs from Category 135. The 1,777 rfMRI IDPs consisted of 76 activity amplitude (node) traits and 1,701 functional connectivity (edge) traits (Category 111). They were parcellation- free and generated by whole brain spatial ICA^1,2,48,49^. Lastly, there were 16 tfMRI IDPs from Category 106. The details of the image acquisition, preprocessing procedures, and quality controls were available in the UKB Brain Imaging Documentation (https://biobank.ctsu.ox.ac.uk/crystal/crystal/docs/brain_mri.pdf). The 41 abdominal MRI IDPs^53,120–128^ were from abdominal organ composition (Category 158), kidney-derived measures (Category 159), liver MRI (Category 126), and abdominal composition (Category 149). For the 82 heart CMR IDPs, the details of the image acquisition, preprocessing procedures, and quality controls were described in previous studies^3,29^. The 46 eye OCT IDPs were all from derived OCT measures^40^ (Category 100079). A list of these IDPs can be found in **Table S1**.

### IGS and dPRS constructions

We performed the following genetic quality controls (QCs) for the set of participants with both IDP and genetic data^37^: 1) removed individuals with missing genotype rate > 0.1; 2) removed variants with missing genotype rate > 0.1; 3) removed variants with minor allele frequency (MAF) < 0.01; and 4) removed variants that failed the Hardy-Weinberg test for equilibrium at 1 × 10^-7^ level. Using individuals of British European ancestry, the GWAS was performed using linear mixed effect models via fastGWA^129^ (average *n* = 34,286 for brain, average *n* = 31,875 for heart, average *n* = 39,830 for abdomen, and average *n* = 54,761 for eye). The adjusting covariates included age (at imaging), age-squared, sex, the interaction between age and sex, the interaction between age-squared and sex, first 40 genetic principal components^130^ (PCs) for all organs; and additionally the estimated total intracranial volume (eTIV), head motion measurements and their squares, brain position measurements and their squares, and volumetric scaling for brain IDPs. Additionally, for BIG-KP regional brain volumes, the total brain volume (TBV) was included as an adjusting covariate to remove global effects^33^. For TBV, the eTIV and volumetric scaling were not included as covariates due to their high linearity. With the GWAS summary statistics as input, we used PRS-CS^55^ and DBSLMM^56^ to obtain the weights for IGS. The hyperparameters of both methods were the default values and/or the automatically tuned values. We then used PLINK^131^ to generate risk scores in testing data by summarizing across genetic variants, weighed by their weights.

The prediction accuracy of IGS was measured by the incremental *R^2^*, which was the additional phenotypic variation that can be explained by the IGS while adjusting for the effects of covariates in a linear regression model. The covariates included age, age- squared, sex, the interaction between age and sex, the interaction between age-squared and sex, and the first 40 genetic PCs. The prediction accuracy was estimated separately in three independent hold-out datasets corresponding to European, Asian, and African ancestries. The European hold-out dataset consisted of UKB individuals of British or non- British European ancestry with IDP data and unrelated to the IGS training set used to generate IDP GWAS (average *n* = 4,541). The Asian hold-out dataset consisted of UKB individuals of Bangladeshi, Chinese, Indian, or Pakistani ancestry with IDP data (average *n* = 460). The African hold-out dataset consisted of UKB individuals of African or Caribbean ancestry with IDP data (average *n* = 252).

We also used PRS-CS^55^ to construct dPRS. Out of the 110 phecode-based diseases that had IGS stratification signals on UKB (**Tables S10-S11**), we mapped 105 diseases to 75 FinnGen R9 disease endpoints that had publicly available GWAS summary statistics (**Table S14**). We used the FinnGen-phecode mapping in Sun et al^97^ and disease description from the FinnGen release R9. For the bipolar disorder analysis on AOU, we used GWAS summary statistics from the Psychiatric Genomics Consortium^132^ to construct BD dPRS.

### IGS-phenotype associations

We used a discovery-replication design to examine associations between IGS and phenotypes in UKB participants without brain or body IDPs. We randomly selected 70% of UKB unrelated British European individuals (average *n* = 202,893) as the discovery dataset for IGS-phenotype associations, while the remaining 30% of UKB unrelated British European individuals, all UKB unrelated non-British European individuals, and all unrelated non-European individuals (average *n* = 129,333) were used as the replication dataset. We treated the values greater than five times the median absolute deviation from the median as outliers and removed these values. A total of 189 UKB phenotypes were tested, which represented a wide range of traits from various domains. Specifically, the 189 UKB phenotypes included 24 mental health traits (Category 100060), 5 cognitive traits (Category 100026), 12 physical activity traits (Category 100054), 6 electronic device use traits (Category 100053), 8 sun exposure traits (Category 100055), 3 sexual factor traits (Category 100056), 3 social support traits (Category 100061), 21 diet traits (Category 100052), 9 alcohol drinking traits (Category 100051), 6 smoking traits (Category 100058), 34 blood biochemistry biomarkers (Category 17518), 3 blood pressure traits (Category 100011), 3 spirometry traits (Category 100020), 17 early life factors (Categories 135, 100033 and 100072), 9 greenspace and coastal proximity (Category 151), 2 hand grip strength (Category 100019), 13 residential air pollution traits (Category 114), 5 residential noise pollution traits (Category 115), 2 body composition traits by impedance (Category 100009), 3 female specific factors (Category 100069), and 1 education trait (Category 100063) (**Table S6**).

Association testing was then conducted to examine the linear relationship between the 4,375 IDP-derived IGS and the 189 UKB phenotypes. We adjusted for the same set of covariates separately in the discovery set and the replication set, including age, age-squared, sex, the interaction between age and sex, the interaction between age-squared and sex, and 40 genetic PCs. Specifically, we regressed the IDP-derived IGS onto the UKB phenotypes and calculated *P*-values using a two-sided t-test. We prioritized the results that met the following three criteria: 1) significant at 0.05 level after Bonferroni correction in the discovery dataset; 2) significant at a nominal significance level of 0.05 in the replication dataset; and 3) had regression coefficients with matching directions in both the discovery and replication datasets.

### Disease risk stratifications

We used IGS to stratify 871 phecode-based^68,69^ diseases (**Table S5**) separately on the UKB discovery cohort and the UKB replication cohort. The UKB discovery cohort consisted of unrelated British European participants without brain or body IDPs (average *n* = 318,781). The UKB replication cohort consisted of unrelated non-British European participants and unrelated non-European participants, all without brain or body IDPs (average *n* = 48,015). In each cohort, we residualized IGS with the same covariates used in the association analysis, standardized the residualized IGS to mean 0 and standard deviation 1, sorted all participants according to the value of the standardized IGS, and split participants into three groups: the group of smallest 10% IGS, the group of largest 10% IGS, and the group of middle 80% IGS. The group of the smallest 10% IGS was referred to as the lower 10% tail, and the group of the largest 10% IGS was referred to as the upper 10% tail. For each disease, the number of disease cases was counted, and a chi-squared test of three levels of IGS-based groups (the smallest 10%, middle 80%, and the largest 10%) and two levels of disease status (case/control) was conducted. The original IGS tail case ratios between the two 10% tails were computed for all IGS-disease pairs and were reported in supplementary tables (**Tables S10-S11**), which were defined as (the number of patients in IGS upper 10% tail) / (the number of patients in IGS lower 10% tail). In **Figures 2** and **S9**, we visualized the maximum of the original IGS tail case ratio and its inverse, that is, a number always greater than or equal to 1. We also only visualized the IGS-disease pairs whose IGS tails contained above-average disease patients. That is, one or both of the two 10% tails of IGS contained at least 10% of all cases on the UKB discovery cohort. In total, 6 pairs between brain IGS and brain/mental disorders, 3 pairs between abdominal IGS and genitourinary/digestive diseases, 3 pairs between heart IGS and circulatory system diseases, and 2 pairs between brain/abdominal/eye IGS and circulatory systems diseases were not visualized because of this.

For Alzheimer’s disease/dementia analysis on AOU, unlike UKB age distribution (**Fig. S15A**), AOU had a bimodal distribution (**Fig. S15B**), and thus we restricted our AOU analysis to participants of age between 60 and 90 (at the age of incidence for patients and at the time of survey otherwise), and only included those self-identified as European (‘White’) (*n* = 59,261, 26,881 males at birth). For the Alzheimer’s disease/dementia patients, we used age at incidence for the analysis in **Figure 4C**, and age at survey for the controls. Due to the lower disease percentage in AOU, we used both Alzheimer’s disease (ICD-10 code: G30) and dementia (ICD-10 code: F01-F03) for cases, and the rest as controls. For multiple sclerosis analysis on AOU, we used all participants self-identified as European (*n* = 128,515, 51,390 males at birth, age range = [17, 113]) and all participants self-identified as African (‘Black’), or Hispanic, or Asian(African *n* = 50,139, 21,684 males at birth, age range = [17, 104]; Hispanic *n* = 41,575, 13,461 males at birth, age range = [17, 100]; Asian *n* = 7,603, 3,066 males at birth, age range = [17, 100]). For bipolar disorder analysis on AOU, we used all participants self-identified as European. Multiple sclerosis patients were those diagnosed with ICD-10 G35, and bipolar disorder patients were those diagnosed with ICD-10 F31. Using PLINK, we developed IGS and dPRS using the Allele Count/Allele Frequency threshold genotyping data prepared by AOU and the UKB weights generated from PRS-CS. We removed the effects of age, sex and 16 genetic PCs from IGS and dPRS, where the genetic PCs were prepared by AOU as well^133^.

For ADNI, we used participants of age at least 60 (*n* = 1,116, 656 males, age range = [60, 91]), the participants whose final diagnosis was dementia were used as cases and the rest as controls (dementia percentage = 42.92%). After performing the standard genetic QCs^33^, we developed the IGS and dPRS using UKB-derived weights and removed the effects of age and sex .

### AUC analysis

To measure the contribution from a single IGS to disease prediction, we trained the following logistic regression models: (1) a baseline model that predicts disease status using age and sex as predictors, and (2) an IGS model that predicts disease status using age, sex, and one IGS. We constructed an IGS model for each IGS-disease pair that had significant stratification (**Tables S12-S13**), a total of 919 IGS models. The increment in the AUC of the IGS model compared with the baseline model is the AUC gain by including a single IGS and visualized in **Figures 3A-C and S9**. DeLong test^96^ was used to test for the difference in AUC between the baseline model and the IGS model. All AUC was obtained using case-control matched individuals. For each disease, each patient was matched with five participants free of this disease^134^, of same age and sex, and of British European ancestry.

To measure the contribution from IGS when dPRS was included, we considered two logistic regression models for disease status prediction: (1) a baseline-dPRS model that included age, sex, and the corresponding dPRS, and (2) an IGS-dPRS model that included age, sex, dPRS, and IGS that marginally stratified this disease. The IGS-dPRS model was selected from the better one between using all stratifying IGS and using only the IGS with the largest stratification tail case ratio. The difference between the AUC of the IGS-dPRS model and the AUC of the baseline-dPRS model was the measure of IGS contribution to disease prediction given dPRS. We averaged the AUC across repeated 100 runs with two-fold cross-validation. The increment in the average AUC of the IGS-dPRS model compared with the average AUC of the baseline-dPRS model is the AUC gain visualized in **Figure 3D**. Wilcoxon rank sum test was used to test for the difference in AUC between the baseline- dPRS model and the IGS-dPRS model across all cross-validation replications.

### IGS burden scores

The brain IGS for Alzheimer’s disease stratification on UKB cohorts was selected according to four criteria below: (1) on the UKB discovery cohort, the IGS stratified disease cases so that the chi-squared test was significant after controlling the FDR at a 5% level; (2) the IGS had one of its two 10% tails containing at least 10% of all disease cases of UKB discovery cohort; (3) the IGS had one of its two 10% tails containing at most 10% of all disease cases of UKB discovery cohort; and (4) the IGS tail case ratio direction was consistent between the UKB discovery cohort and the UKB replication cohort (that is, the 10% tail that contained more disease cases was consistent on both cohorts). There were 55 IGS that satisfied these conditions and were selected for Alzheimer’s disease stratification (**Table S16**). In **Figures 4A-4B**, **4E-4F** and **5**, within each group (defined by age, or by dPRS, or by *APOE* status, or by both *APOE* status and dPRS), brain IGS were selected with the following two criteria: (1) the IGS was selected for disease stratification on the entire cohort (that is, among the 55 selected IGS above); and (2) the 10% IGS tail in the group that contained more cases was also the 10% IGS tail that had contained more cases on the entire cohort (that is, the IGS high-risk tail that contained more disease cases remained consistent in this age/dPRS/*APOE*-defined group).

IGS burden score of a participant was the number of “IGS high-risk tails” a participant was in. The “IGS high-risk tail” for IGS burden score was the 20% top tail of an IGS that contained more patients compared to the lower 20% tail. We constructed an IGS burden score by counting the number of IGS high-risk tails a subject was in, across the selected IGS. For example, if a subject was ranked in the top 20% for five out of the selected brain IGS, they would be assigned an IGS burden score of five. As IGS burden score is an integer as small as zero, for each group, we selected the “Low IGS burden score” to be the smallest nonnegative integer values that corresponded to around 20% of all participants in this group, and the “Higher IGS burden score” to be the largest integer values that corresponded to around 20% of all participants in this group. Within a group, the participants with IGS burden score among the “Lower IGS burden score” values were categorized as “Lower IGS burden score”, and likewise, those with IGS burden score among the “Higher IGS burden score” values were categorized as “Higher IGS burden score”. Due to smaller sample size and higher dementia percentage in ADNI, ADNI IGS burden score used 10% tail for “IGS high-risk tail”. Besides, ADNI had “Lower IGS burden score” cutoff at 10% for age analysis (**Fig. 4D**).

Similar to UKB analysis, we grouped AOU participants of European ancestry into different groups (by age as in **Fig. 4C** or by dPRS as in **Fig. 4G**). For each stratum, we again calculated the IGS tail case ratio for the 55 selected IGS and only kept the IGS that had case ratio direction consistent with the UKB discovery cohort, and categorized participants inside each stratum into “Lower IGS burden score” and “Higher IGS burden score”. Similar analysis was performed for ADNI participants (**Figs. 4D** and **4H**).

For multiple sclerosis stratification on UKB and AOU, similar to Alzheimer’s disease, we applied the following criterion to select the brain IGS for IGS burden score: (1) on the UKB discovery cohort, the IGS stratified multiple sclerosis cases so that the chi-squared test was significant after controlling the FDR at a 5% level; (2) the IGS had one of its two 10% tails containing at least 10% of all multiple sclerosis cases of UKB discovery cohort; (3) the IGS had one of its two 10% tails containing at most 10% of all multiple sclerosis cases of UKB discovery cohort; and (4) the IGS tail case ratio direction was consistent between the UKB discovery cohort and the UKB replication cohort (that is, the 10% tail that contained more cases was consistent on both cohorts). There were 25 IGS that satisfied these conditions and were selected for IGS burden score of multiple sclerosis (**Table S17**), and then stratification within group defined by multiple sclerosis dPRS were carried out using IGS burden score, similar to Alzheimer’s disease analysis described above.

### *APOE* status

We extracted *APOE* genotypes for UKB participants based on the genetic variants rs429358 and rs7412. Following prior studies on *APOE*^135^, we considered five common *APOE* genotypes (ε4ε4, ε3ε4, ε3ε3, ε2ε3 and ε2ε2) and mapped the alleles of rs429358 and rs7412 as follows: rs429358 CC and rs7412 CC to ε4ε4, rs429358 TC and rs7412 CC to ε3ε4, rs429358 TT and rs7412 CC to ε3ε3, rs429358 TT and rs7412 TC to ε2ε3, and rs429358 TT and rs7412 TT to ε2ε2. Here ε4ε4 are ε4 homozygotes, ε3ε4 are ε4 heterozygotes, ε3ε3 are ε3 homozygotes, and ε2ε2 and ε2ε3 are ε2 carriers. We also combined ε4 homozygotes and ε4 heterozygotes as “ε4 carriers” in **Figures 5A and 5E**.

### Code availability

We made use of publicly available software and tools. The list of genetic variants and their weights used for the construction of IGS for UKB brain and body IDPs are available at https://github.com/xcyang17/IPRS_UKB.

### Data availability

The individual-level data used in this study can be obtained from https://www.ukbiobank.ac.uk/ and http://adni.loni.usc.edu/data-samples/. The disease GWAS data can be downloaded from https://www.finngen.fi/en/access_results and https://figshare.com/articles/dataset/bip2021_noUKBB/22564402. Our IGS data resources can be downloaded at Zenodo (https://doi.org/10.5281/zenodo.7709788).

