## supplementary_info for "Multi-organ imaging-derived polygenic indexes for brain and body health"

**This PDF file includes:**

Supplementary Note
Figs. S1 to S16
Legends for Tables S1 to S18

**Other Supplementary Materials for this manuscript include the following:**

Tables S1 to S18 (.xlsx) (available in a zip file)

### Supplementary Note

#### Brain IGS-phenotype associations.

We provided examples between brain IGS and complex traits that were consistent between BIG-KP and UKB-Oxford IDPs. In total, 27 out of the 34 blood biomarkers were found associated with the IGS of BIG-KP brain IDPs, including biomarkers closely related to cardiovascular health, such as cholesterol, LDL, and triglycerides (UKB Data-Field 30690, 30780, and 30870) ($|\beta| > 0.0104$, *P* range = [$7.79 \times 10^{-47}$, $8.07 \times 10^{-7}$]). For example, LDL was found positively associated with IGS of axial diffusivity of various brain regions ($\beta > 0.0105$, *P* range = [$4.06 \times 10^{-22}$, $7.06 \times 10^{-7}$]), consistent with prior findings on the relationship between LDL and axial diffusivity[1]. Similarly, among the IGS of UKB-Oxford brain IDPs, 28 out of the 34 blood biomarkers had significant associations, and all associations between axial diffusivity IGS and LDL were positive ($\beta > 0.0118$, *P* range = [$3.56 \times 10^{-27}$, $2.72 \times 10^{-8}$]).

Among the 24 mental health traits, 15 were found associated with the IGS of BIG-KP brain IDPs. All these 15 mental health traits were associated with IGS of BIG-KP regional brain volumes. Nervous feelings, visits to doctors/psychiatrists, and neuroticism score were also widely associated with IGS of BIG-KP DTI parameters, whereas neuroticism score was also associated with BIG-KP Glasser360-based rfMRI IGS ($|\beta| > 0.0104$, *P* range = [$1.25 \times 10^{-16}$, $8.63 \times 10^{-7}$]). For example, the IGS of FA of the body and genus of the corpus callosum was negatively associated with neuroticism score ($\beta$=-0.0135 and -0.0139; $P = 8.43 \times 10^{-9}$ and $2.67 \times 10^{-9}$, respectively). Imaging-based studies showed that subjects with high anxiety or depression tend to have decreased FA in the corpus callosum[2,3]. The same 15 mental health traits were found associated with the IGS of UKB-Oxford brain IDPs, and consistent patterns were observed ($|\beta| > 0.0113$, *P* range = [$1.65 \times 10^{-21}$, $6.75 \times 10^{-8}$]), including the significant association between the 15 mental health traits with sMRI IGS, the wide association between the aforementioned traits with dMRI IGS, and the association between neuroticism score with ICA-based rfMRI IGS. For example, the IGS of the BIG-KP total brain volume and the IGS of the UKB-Oxford whole brain volume (Data-Field 26521) were both negatively associated with neuroticism score (BIG-KP $|\beta|$ = -0.0178, $P = 1.82 \times 10^{-14}$; UKB-Oxford $|\beta|$ = -0.0215, $P = 3.98 \times 10^{-20}$), consistent with the known links between neuroticism and reduced brain volume from imaging studies[4,5].

The 1,439 sMRI traits in the UKB-Oxford consisted of multiple subcategories, including regional volumes, cortical areas, cortical grey-white contrast, cortical thickness, regional and tissue intensity, regional T2*, and white matter hyperintensity volume (**Table S1**). Therefore, the IGS of some sMRI subcategories exclusive to UKB-Oxford revealed additional associations that were not detected by BIG-KP IGS. For example, playing computer games (Data-Field 2237), a possibly addictive behavior, was found negatively associated with the area of the left inferior temporal (Data-Field 27149, $\beta$ = -0.0115, $P = 4.35 \times 10^{-8}$). It was reported that young male adults playing internet video games had smaller inferior temporal gyri[6].

#### Body IGS-phenotype associations.

We provided examples between body IGS and complex traits. Among abdominal MRI-based IGS, we found wide associations between liver MRI IGS and blood biochemistry biomarkers, such as albumin, direct bilirubin, gamma-glutamyl transferase, alanine transaminase (ALT), and aspartate aminotransferase (AST) (Data-Field 30600, 30660, 30730, 30620, and 30650, respectively; ($\beta$ > 0.0137, $P$ range = [$5.96 \times 10^{-295}$, $1.74 \times 10^{-9}$]). For example, we found both AST and ALT positively associated with the IGS of the fat referenced liver proton density fat fraction (Data-Field 24352). Previous MRI studies showed liver fat fraction was positively correlated with AST and ALT[7], and ALT has been a commonly used biomarker to evaluate fatty liver disease progression[8]. As for heart CMR-based IGS, associations with blood biochemistry biomarkers, blood pressure, body impedance, and spirometry were the majority. For example, descending aortic distensibility was found negatively correlated with both diastolic blood pressure (Data-Field 4079) and systolic blood pressure (Data-Field 4080), consistent with multiple MRI studies that found a negative correlation between blood pressure and aortic distensibility[9,10], and aortic distensibility is a known factor mechanically affecting blood pressure[11]. For eye OCT-based IGS, we found the IGS of the thickness of the INL of both eyes was positively associated with diastolic blood pressure and systolic blood pressure ($\beta$ > 0.0112, $P$ range = [$7.58 \times 10^{-9}$, $5.95 \times 10^{-8}$], Data-Field 28502 and 28503), and a population-based study showed that higher blood pressure was correlated with thicker INL[12].

**Marginal stratification of Alzheimer's disease by brain IGS.**

The IGS of Net25_Node9, an ICA-based rfMRI activity trait[13] for the visual network (mainly within lingual, calcarine, superior occipital regions), contained a 2.46-fold of Alzheimer's disease cases in its lower 10 % tail when compared with its upper 10% tail ($P = 6.35 \times 10^{-37}$). A further stratification analysis across all values of Net25_Node9 IGS showed the disease percentage monotonely decreased as Net25_Node9 IGS increased on the UKB discovery cohort, and relatively monotonely on the UKB replication cohort (**Fig. S8** and **Table S16**).

**AUC analysis.**

We listed a few diseases with varying AUC gain from IGS when disease polygenic risk score (dPRS) was already included. Diseases that had uninformative dPRS and large AUC gain from IGS: cerebral degeneration, unspecified (**Fig. 3E**), and nephritis and nephropathy without mention of glomerulonephritis (**Fig. S11H**). Diseases that had informative dPRS and moderate AUC gain from IGS: delirium due to conditions classified elsewhere (**Fig. 3F**), aortic aneurysm (**Fig. 3G**), myopia (**Fig. S11C**), primary/intrinsic cardiomyopathies (**Fig. S11D**), chronic renal failure (**Fig. S11I**), other chronic nonalcoholic liver disease (**Fig. S11F**), and portal hypertension (**Fig. S11G**). Diseases that had very informative dPRS and small AUC gain from IGS: Alzheimer's disease (**Fig. 3H**), multiple sclerosis (**Fig. S11A**), and open−angle glaucoma (**Fig. S11B**). Lastly, disease that had very informative dPRS and moderate AUC gain from IGS: intestinal malabsorption (non−celiac) (**Fig. S11E**).

**Bipolar disorder stratification on AOU.**

We conducted stratification analysis of bipolar disorder (BD) on AOU European cohort and stratified BD using 383 brain IGS and 169 body IGS. After controlling FDR at 5% level, one brain IGS (total brain volume) significantly stratified BD (IGS tail case ratio = 0.82, $P = 4.35 \times 10^{-3}$). The IGS of total brain volume stratified BD risk across all values (**Fig. S16A** and **Table S18**). In **Figure S16B**, within BD dPRS-defined strata, those of bottom 20% IGS (“Lower IGS”) always had higher BD risk than those of top 20% IGS (“Higher IGS”), except for the stratum of lowest 20% dPRS. The

BD dPRS was constructed using a UKB-independent GWAS summary statistics from the Psychiatric Genomics Consortium[14] and stratified BD patients well (**Fig. S16C** and **Table S18**).

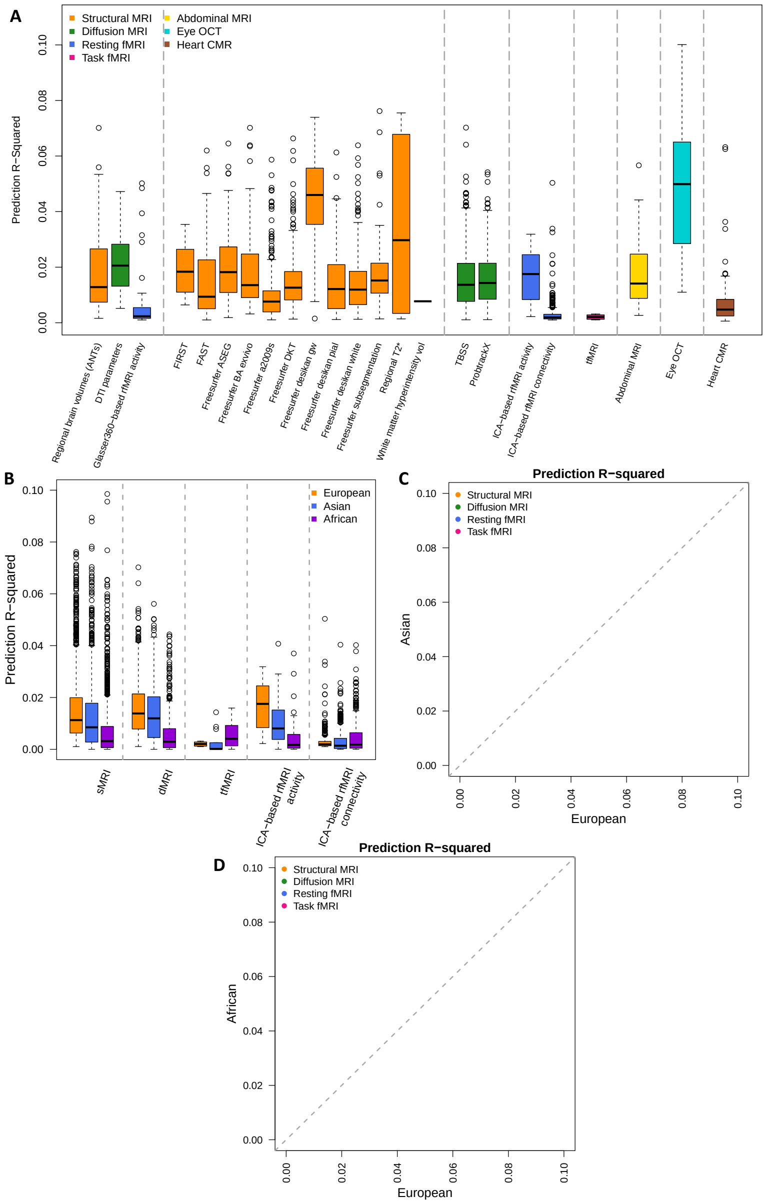


**Fig. S1 Prediction analysis.**

**(A)** The incremental prediction $R^2$ of all brain and body IGS on the UKB European hold-out testing datasets. From left to right, the 8 sections separated by vertical grey dashed lines correspond to the following modalities: BIG-KP IDPs (sMRI, dRMI, and rfMRI), UKB-Oxford sMRI IDPs, UKB-Oxford dMRI IDPs, UKB-Oxford rfMRI IDPs, UKB-Oxford tfMRI IDPs, abdominal MRI IDPs, eye OCT IDPs, and heart CMR IDPs. A complete list of the traits can be found in Table S1. Only IDPs that were significantly predicted by the corresponding IGS after controlling the FDR rate at a 5% level are displayed. **(B)** The incremental prediction $R^2$ of UKB-Oxford brain IGS included in **(A)** on the UKB European, Asian, and African hold-out testing datasets. **(C)** The incremental prediction $R^2$ of UKB-Oxford brain IGS included in **(A)** on the UKB European and Asian testing datasets. **(D)** The incremental prediction $R^2$ of UKB-Oxford brain IGS included in **(A)** on the UKB European and African testing datasets.

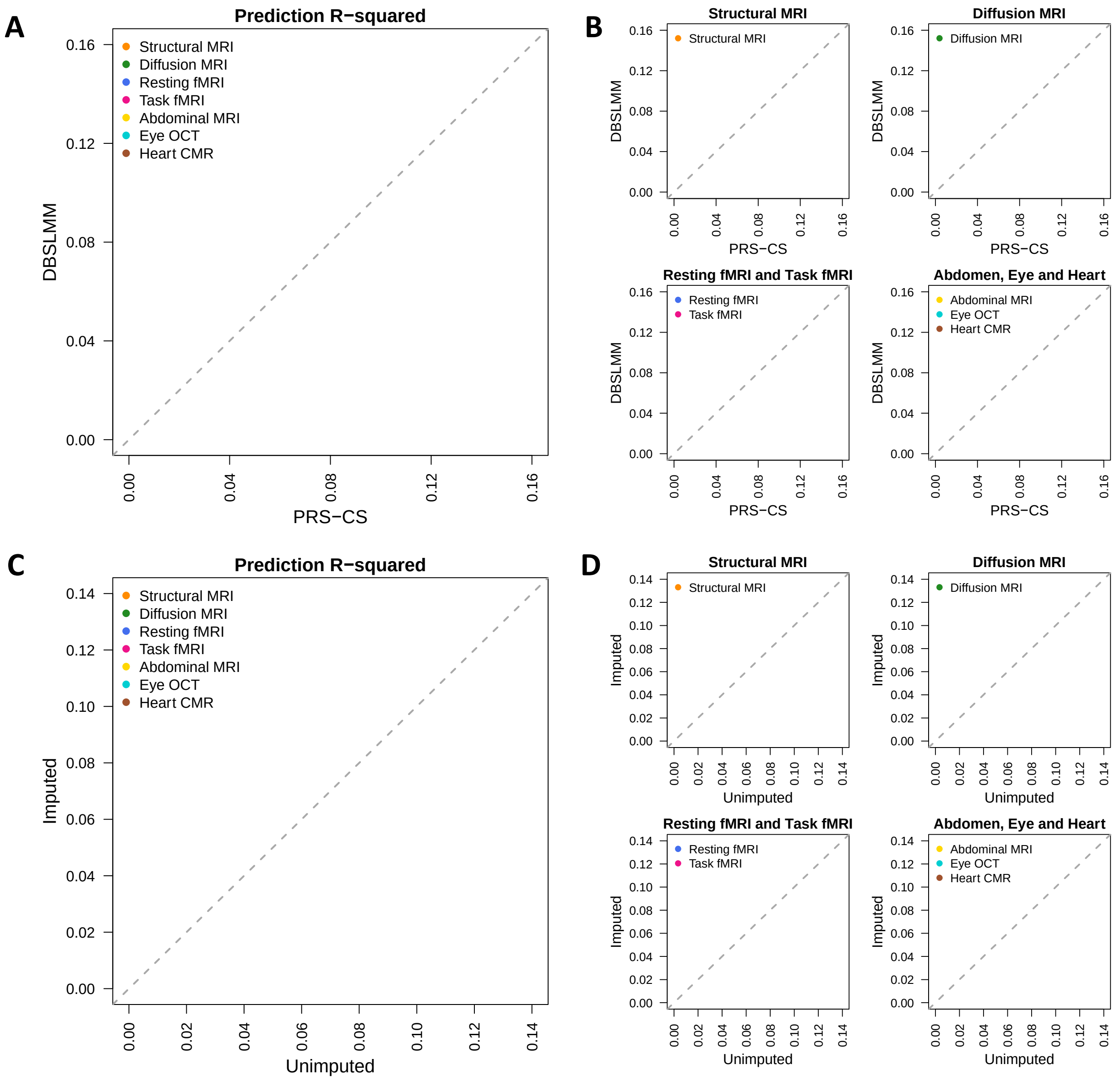


**Fig. S2 Prediction analysis of different IGS methods.**

**(A)** Comparison of incremental prediction $R^2$ of IGS in PRS-CS and DBSLMM. **(B)** Comparison of incremental prediction $R^2$ of IGS in **(A)** split by imaging modalities. **(C)** Comparison of incremental prediction $R^2$ of IGS using unimputed genotyping data and imputed genotyping data, both using PRS-CS. **(D)** Comparison of incremental prediction $R^2$ of IGS in **(C)** split by imaging modalities. The grey dashed line is the 45-degree line. Only IDPs that were significantly predicted by the corresponding PRS-CS IGS on the UKB European hold-out testing dataset after controlling the FDR rate at a 5% level are displayed.

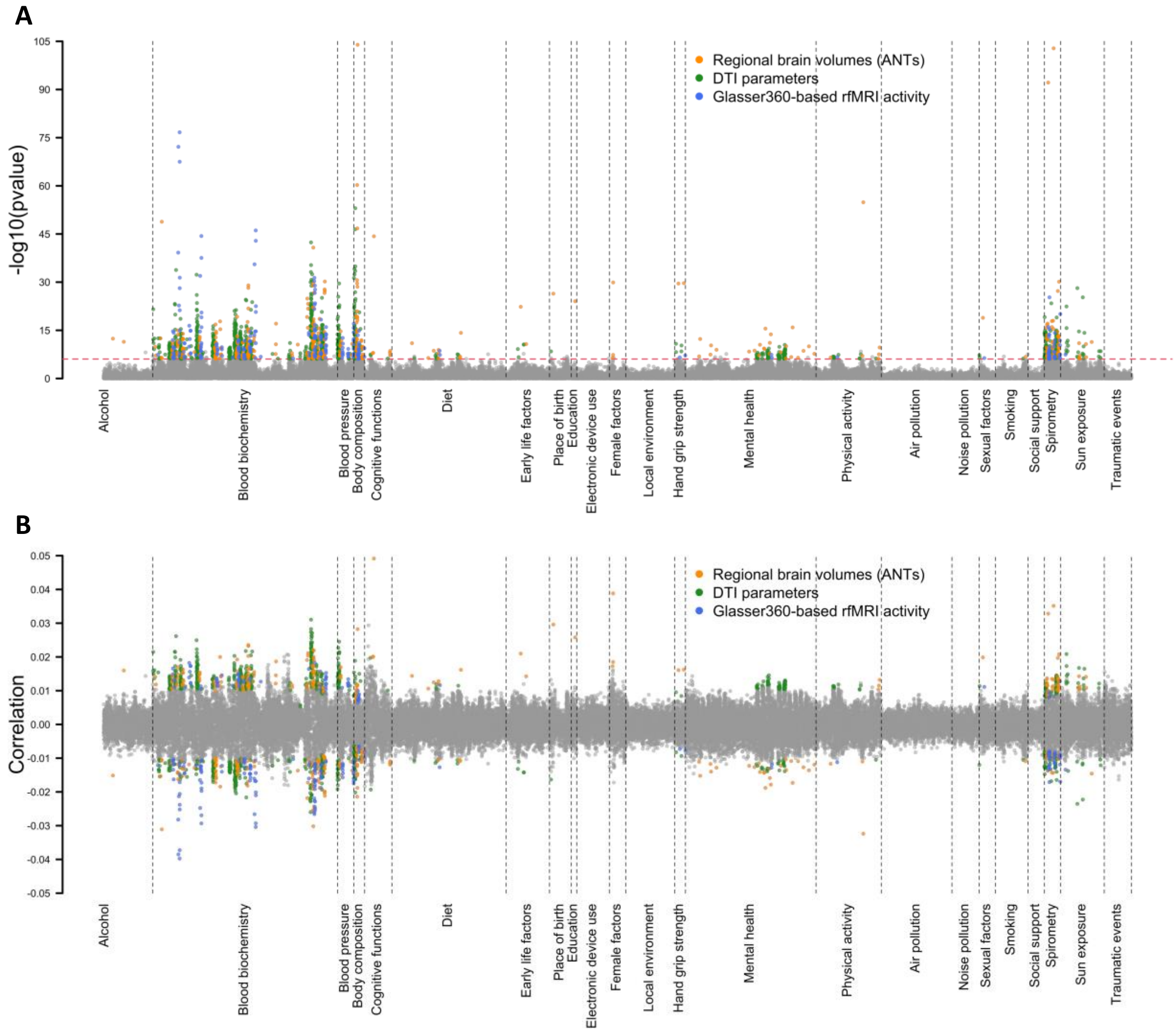


**Fig. S3 BIG-KP brain IGS-phenotype associations that can be replicated in an independent hold-out testing dataset.**

The *P*-values (upper panel) and correlation coefficients (lower panel) between 189 phenotypes and the IGS of 3 modalities of BIG-KP brain MRI traits, including 101 regional brain volumes, 110 DTI parameters, and 90 resting fMRI traits. The Bonferroni-significance level ($P < 8.79 \times 10^{-7}$, horizontal red dashed line in upper panel) coefficients replicated in the independent hold-out sample are highlighted in colors. We label the categories of brain imaging traits with different colors.

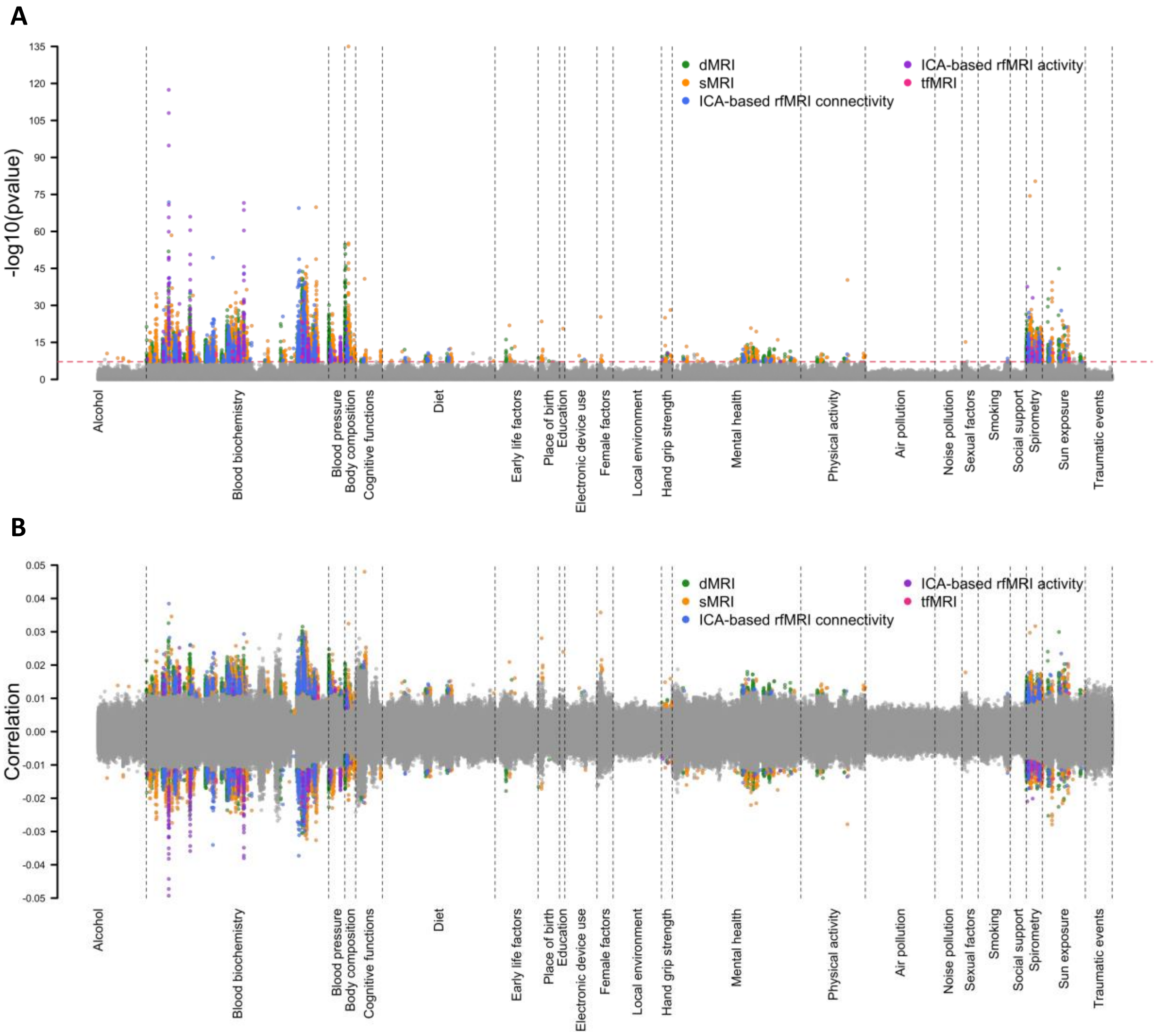


**Fig. S4 UKB-Oxford brain IGS-phenotype associations that can be replicated in an independent hold-out testing dataset.**

The *P*-values (upper panel) and correlation coefficients (lower panel) between 189 phenotypes and the IGS of 5 modalities of UKB-Oxford brain MRI traits, including 657 dMRI, 1,437 sMRI, 1,701 resting fMRI connectivity, 76 resting fMRI activity, and 16 task fMRI traits. The Bonferroni-significance level ($P < 6.77 \times 10^{-8}$, horizontal red dashed line in upper panel) coefficients replicated in the independent hold-out sample are highlighted in colors. We label the categories of brain imaging traits with different colors.

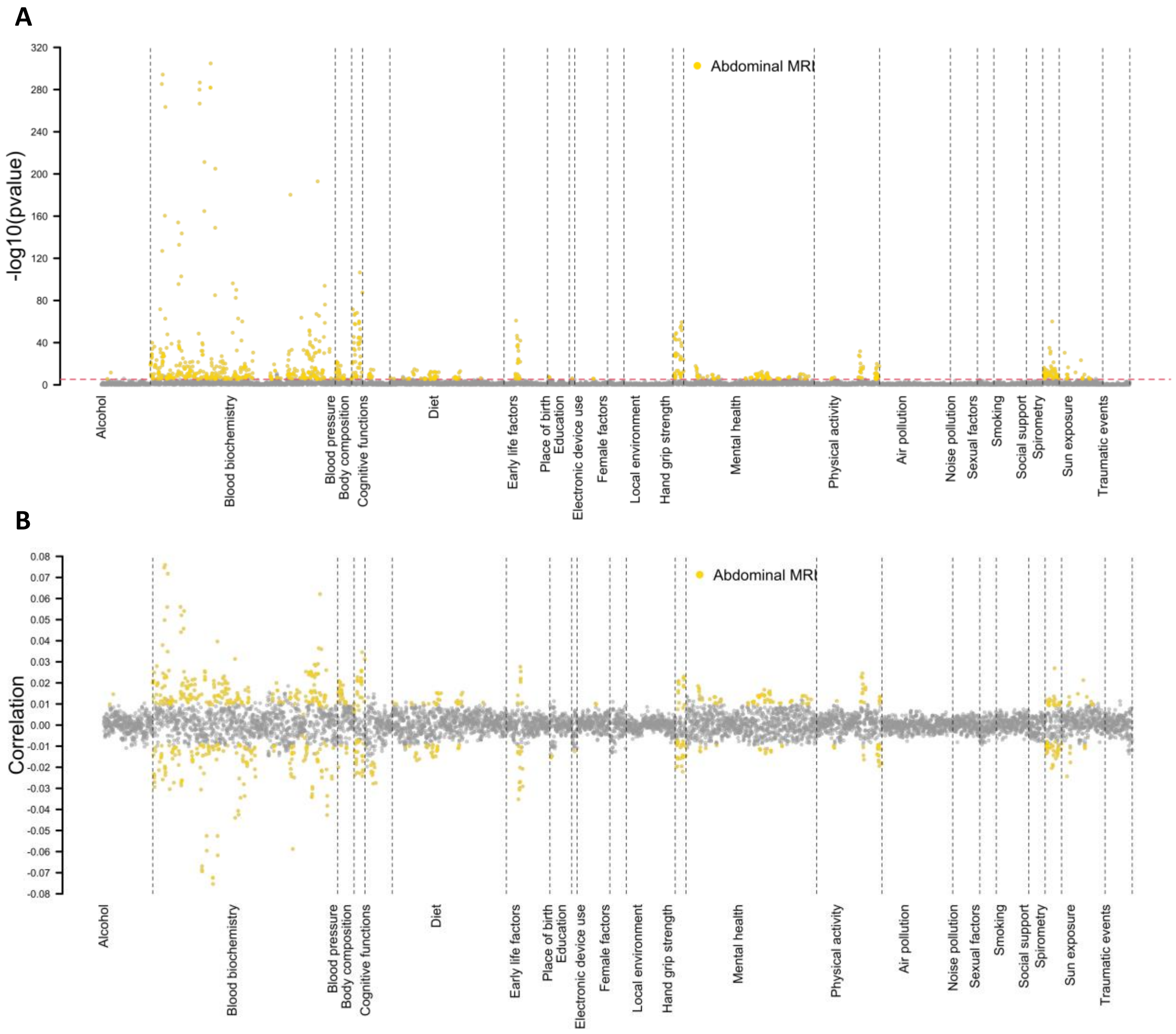


**Fig. S5 Abdominal IGS-phenotype associations that can be replicated in an independent hold-out testing dataset.**

The *P*-values (upper panel) and correlation coefficients (lower panel) between 189 phenotypes and the IGS of 41 abdominal MRI traits. The Bonferroni-significance level ($P < 6.45 \times 10^{-6}$, horizontal red dashed line in upper panel) coefficients replicated in the independent hold-out sample are highlighted in colors. We label the categories of brain imaging traits with different colors.

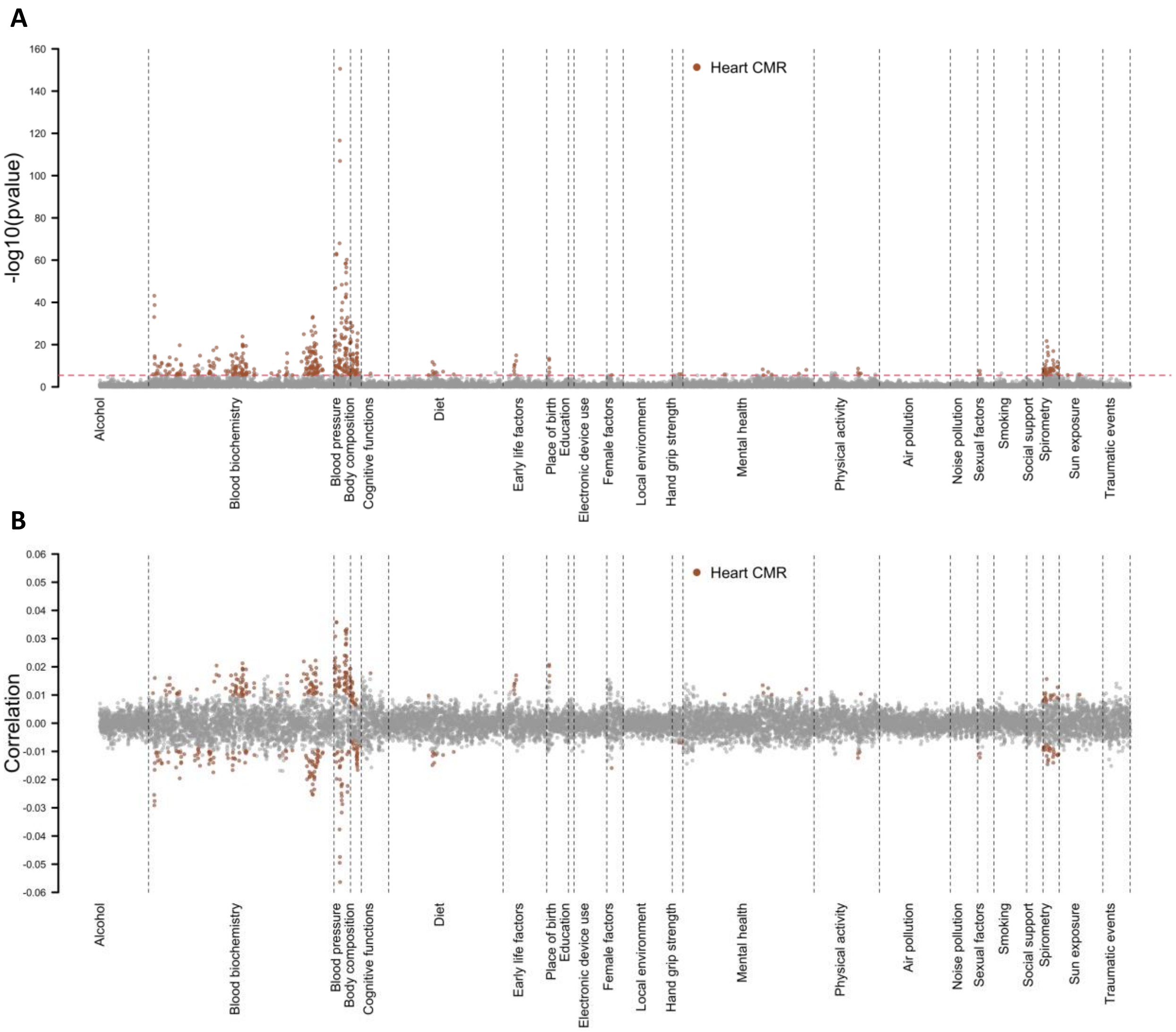


**Fig. S6 Heart IGS-phenotype associations that can be replicated in an independent hold-out testing dataset.**

The *P*-values (upper panel) and correlation coefficients (lower panel) between 189 phenotypes and the IGS of 82 UKB heart CMR traits. The Bonferroni-significance level ($P < 3.23 \times 10^{-6}$, horizontal red dashed line in upper panel) coefficients replicated in the independent hold-out sample are highlighted in colors. We label the categories of brain imaging traits with different colors.

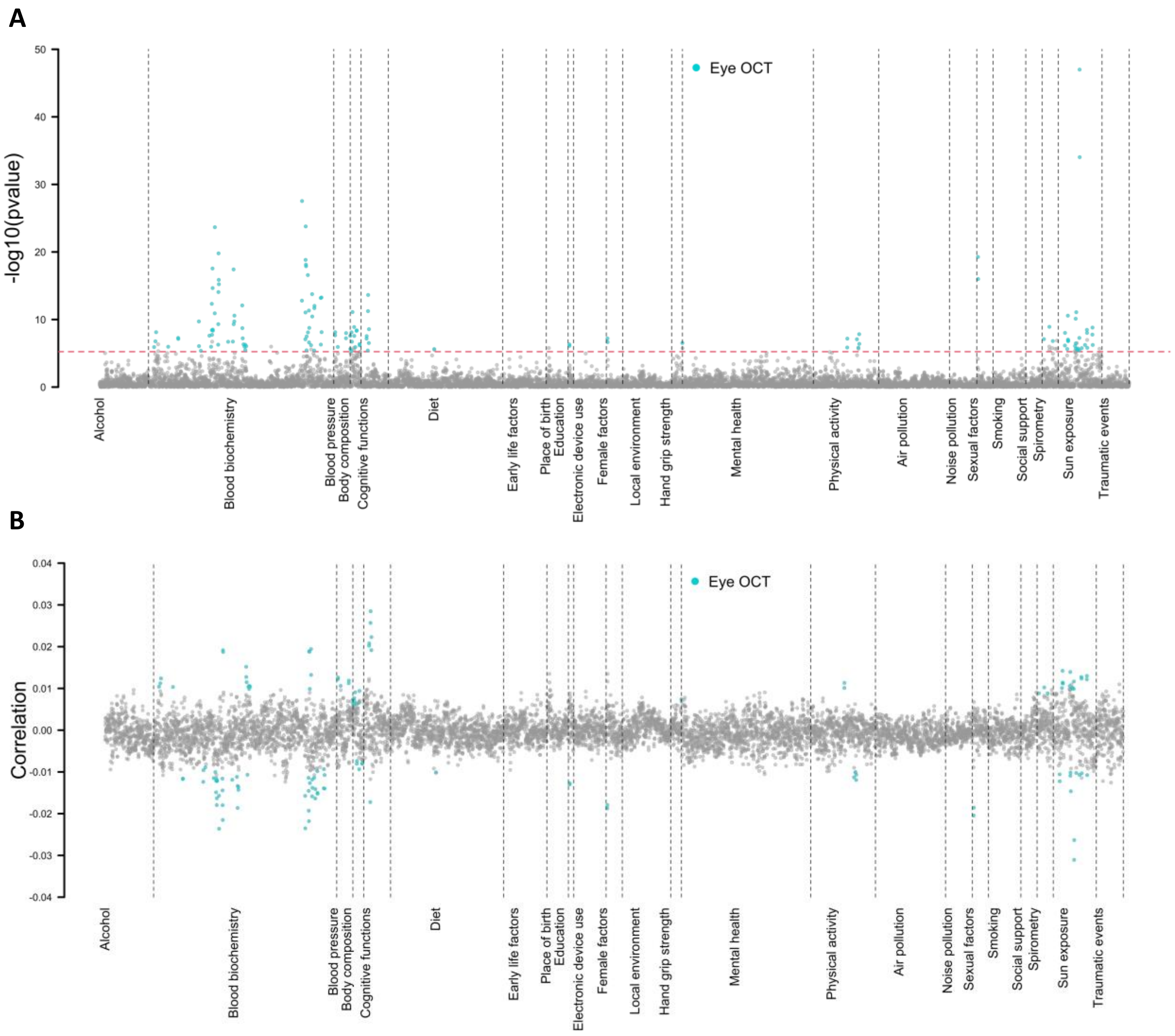


**Fig. S7 Eye IGS-phenotype associations that can be replicated in an independent hold-out testing dataset.**

The *P*-values (upper panel) and correlation coefficients (lower panel) between 189 phenotypes and the IGS of 46 UKB eye OCT traits. The Bonferroni-significance level ($P < 5.75 \times 10^{-6}$, horizontal red dashed line in upper panel) coefficients replicated in the independent hold-out sample are highlighted in colors. We label the categories of brain imaging traits with different colors.

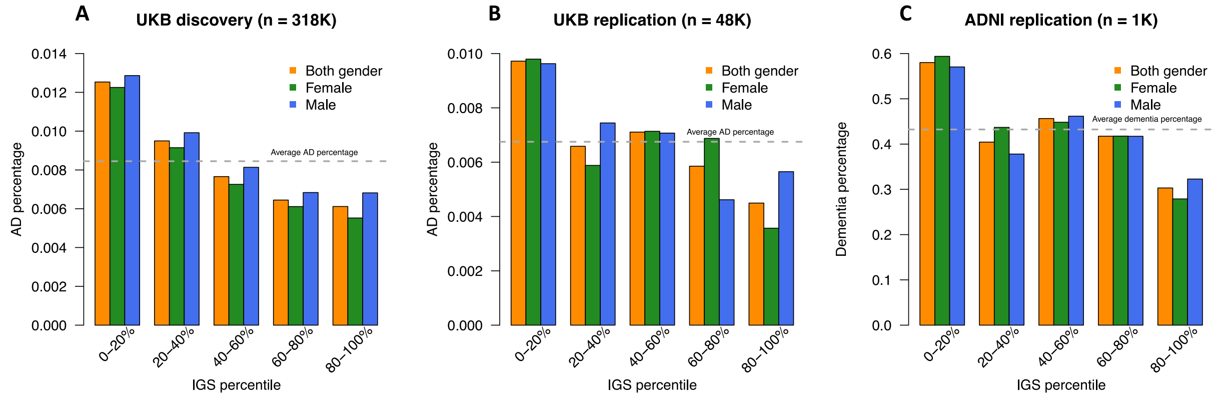


**Fig. S8 Stratification of Alzheimer’s disease by Net25_Node9 IGS.**

For the pair between Alzheimer’s disease (AD) and Net25_Node9, we show the disease percentage (*y*-axis) in quantile-based groups defined by Net25_Node9 IGS (*x*-axis) in the UKB discovery, UKB replication, and Alzheimer's Disease Neuroimaging Initiative (ADNI) cohorts. The horizontal grey dashed line represents the disease percentage for the entire cohort.

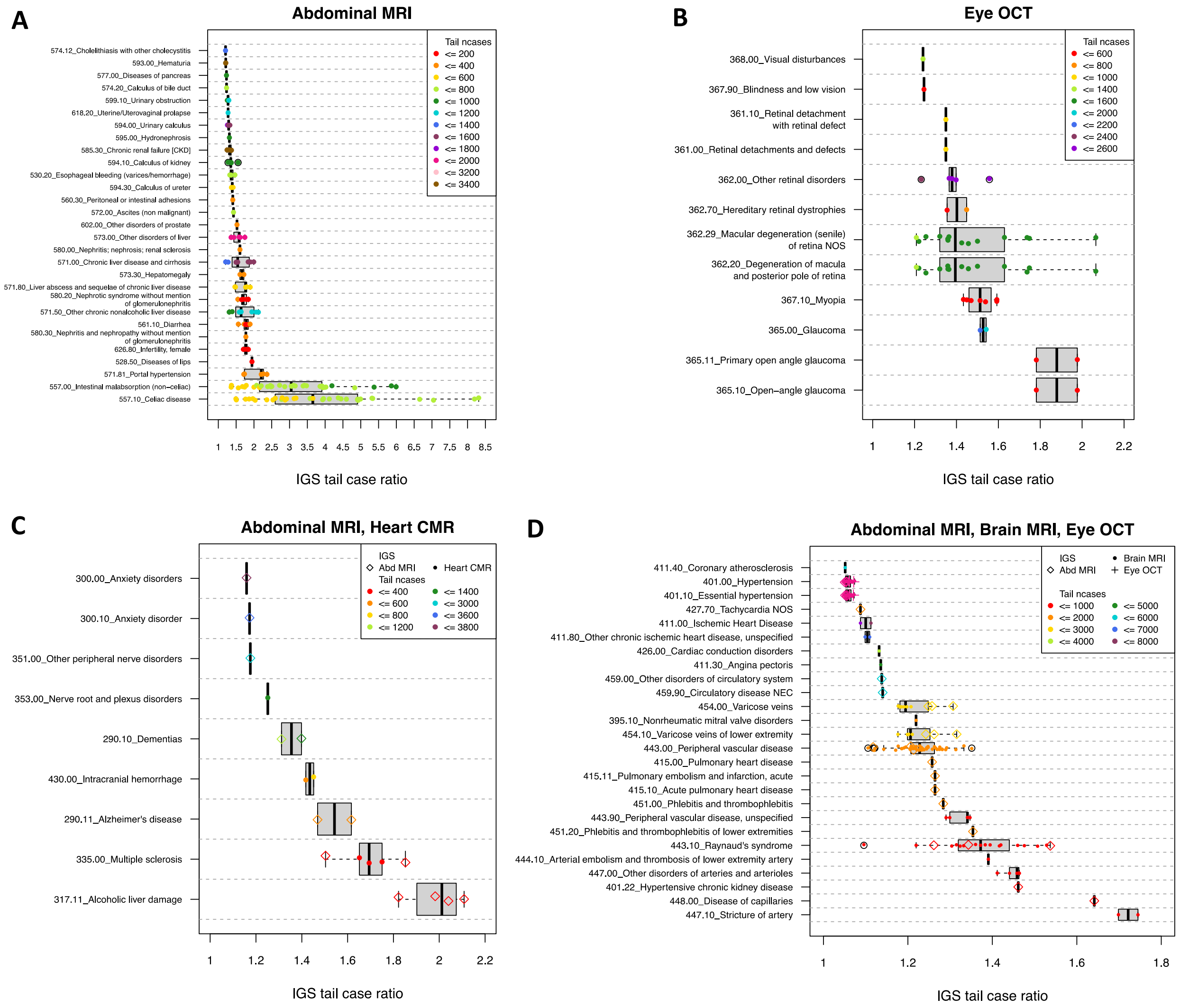


**Fig. S9 IGS stratification of organ-specific disorders and cross-organ disorders.**

**(A)** IGS stratification of 104 digestive system disorders and 100 genitourinary system disorders using 46 abdominal IGS. Each point represents an IGS-disease pair. **(B)** IGS stratification of 38 eye and adnexa disorders using 46 eye IGS. **(C)** IGS stratification of 69 brain and mental disorders using 41 abdominal IGS and 82 heart IGS. **(D)** IGS stratification of 90 circulatory system diseases using 41 abdominal IGS, 383 brain IGS and 46 eye IGS. Only significant IGS-disease pairs after controlling the FDR rate at a 5% level are displayed. In **(A)-(B)**, only IGS-disease pairs with IGS tail case ratio greater than 1.2 are displayed. In **(C)-(D)**, only IGS-disease pairs with IGS tail case ratio greater than 1.05 are displayed. We show the results for UKB discovery cohort.

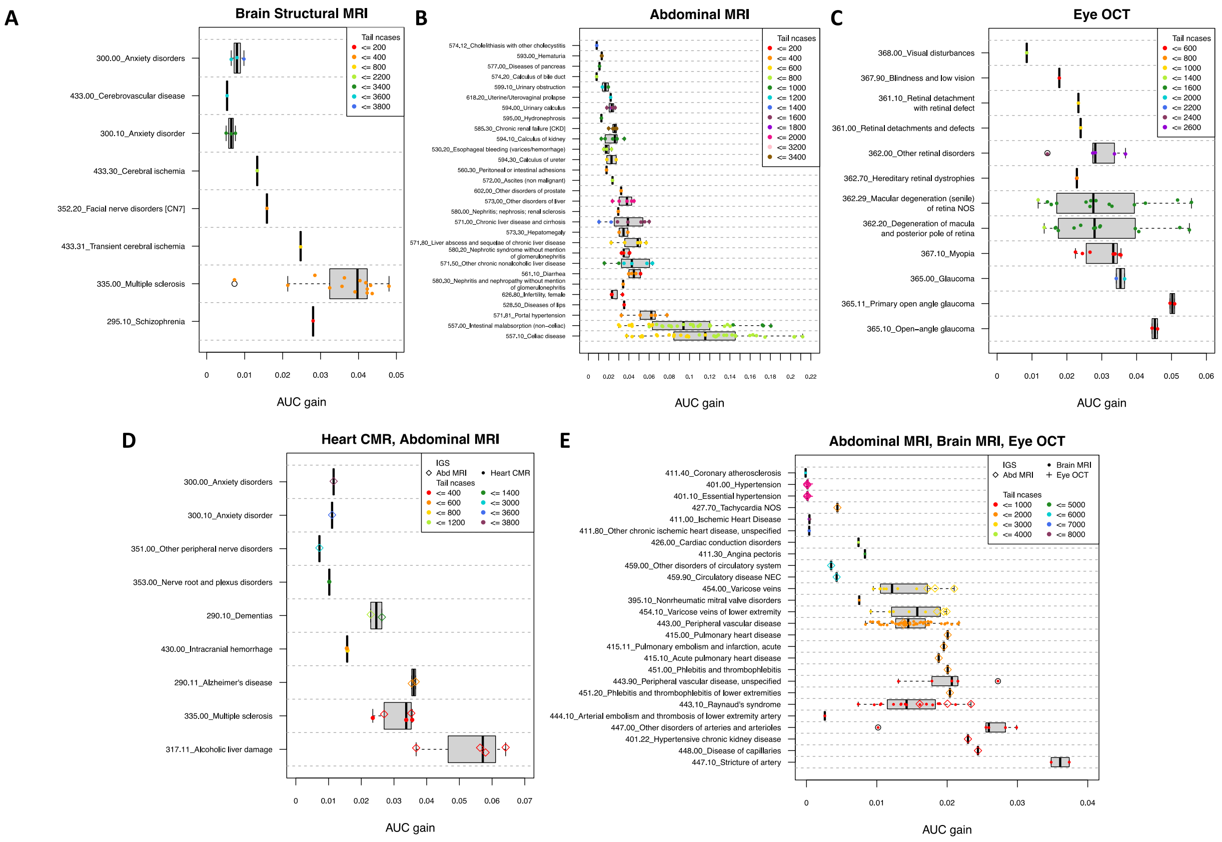


### Fig. S10 AUC gain from stratifying IGS for prediction of organ-specific disorders and cross-organ disorders.

AUC gain by including the stratifying IGS for IGS-disease pairs that were significant on UKB discovery cohort after controlling the FDR rate at a 5% level and replicated on UKB replication cohort. **(A)** AUC gain for significant and replicated IGS-disease pairs between brain structural MRI IGS and brain and mental disorders. **(B)** AUC gain for significant and replicated IGS-disease pairs between abdominal IGS and digestive system disorders / genitourinary system disorders. **(C)** AUC gain for significant and replicated IGS-disease pairs between eye IGS and eye and adnexa disorders. **(D)** AUC gain for significant and replicated IGS-disease pairs between heart IGS and abdominal IGS and brain and mental disorders. **(E)** AUC gain for significant and replicated IGS-disease pairs between abdominal IGS, brain IGS and eye IGS and circulatory system disease. In **(A)**, only IGS-disease pairs with IGS tail case ratio greater than 1.1 are displayed. In **(B)-(C)**, only IGS-disease pairs with IGS tail case ratio greater than 1.2 are displayed. In **(C)-(D)**, only IGS-disease pairs with IGS tail case ratio greater than 1.05 are displayed. We show the results for UKB discovery cohort.

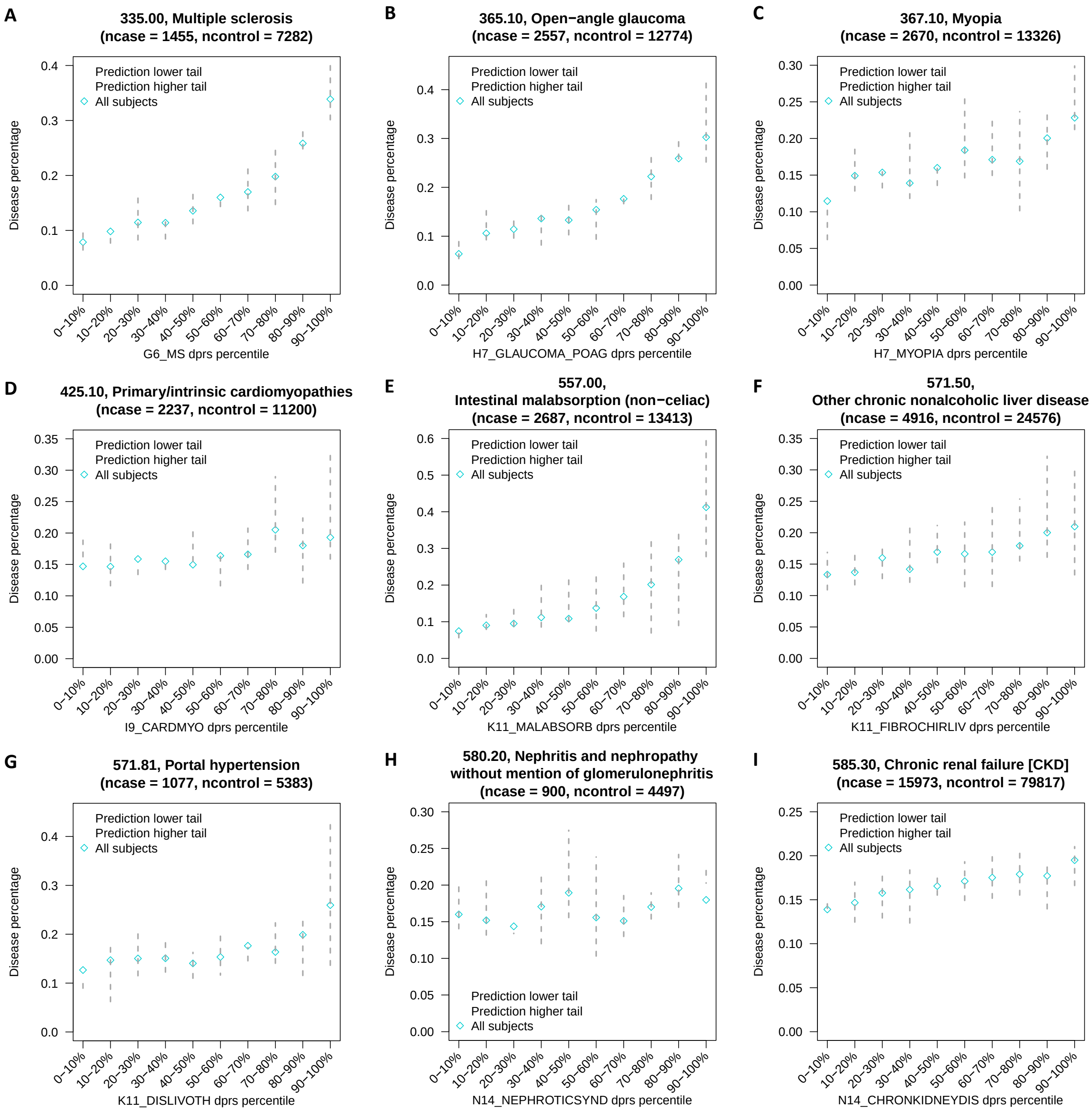


**Fig. S11 IGS stratification of diseases.**

**(A)-(I)** For nine select diseases, for each dPRS quantile group (*x*-axis), we show the disease percentage (*y*-axis) for all participants ("All subjects"), participants in the low 10% tail of the prediction of the IGS-dPRS model of the disease ("Prediction lower tail"), and participants in the high 10% tail of IGS burden scores ("Prediction higher tail"). We show the results for UKB discovery cohort.

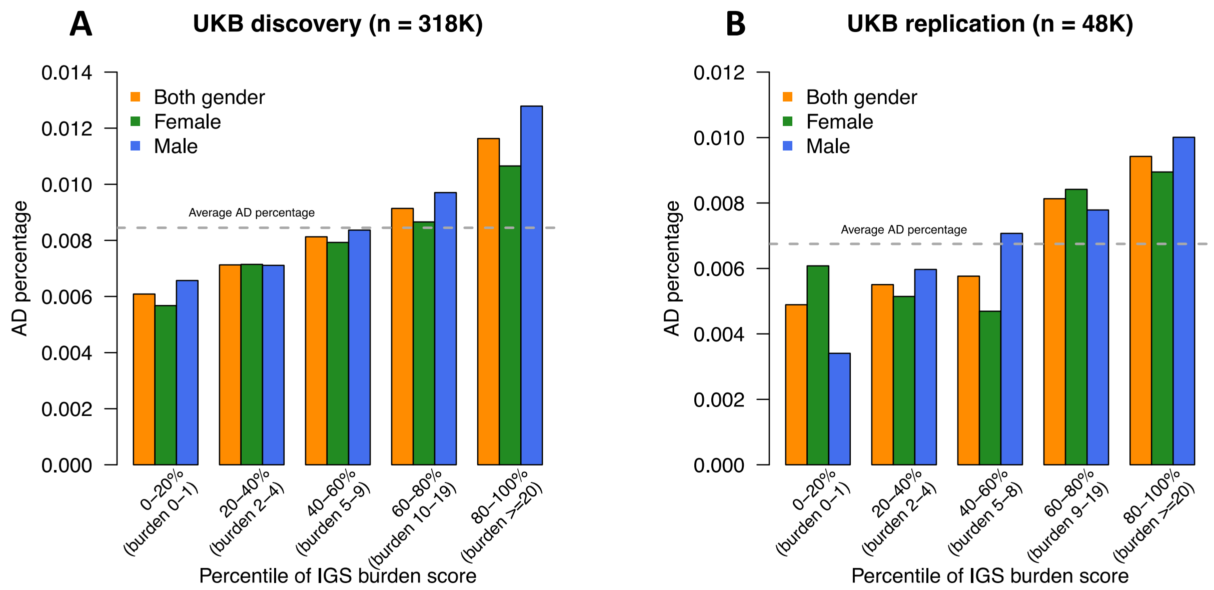


**Fig. S12 IGS burden score stratification of Alzheimer's disease.**

**(A)-(B)** In each IGS burden score quantile group (*x*-axis), we show the Alzheimer's disease (AD) percentage (*y*-axis) for all subjects ("Both gender"), females ("Female"), and males ("Male"). We show the results for UKB discovery and UKB replication cohorts.

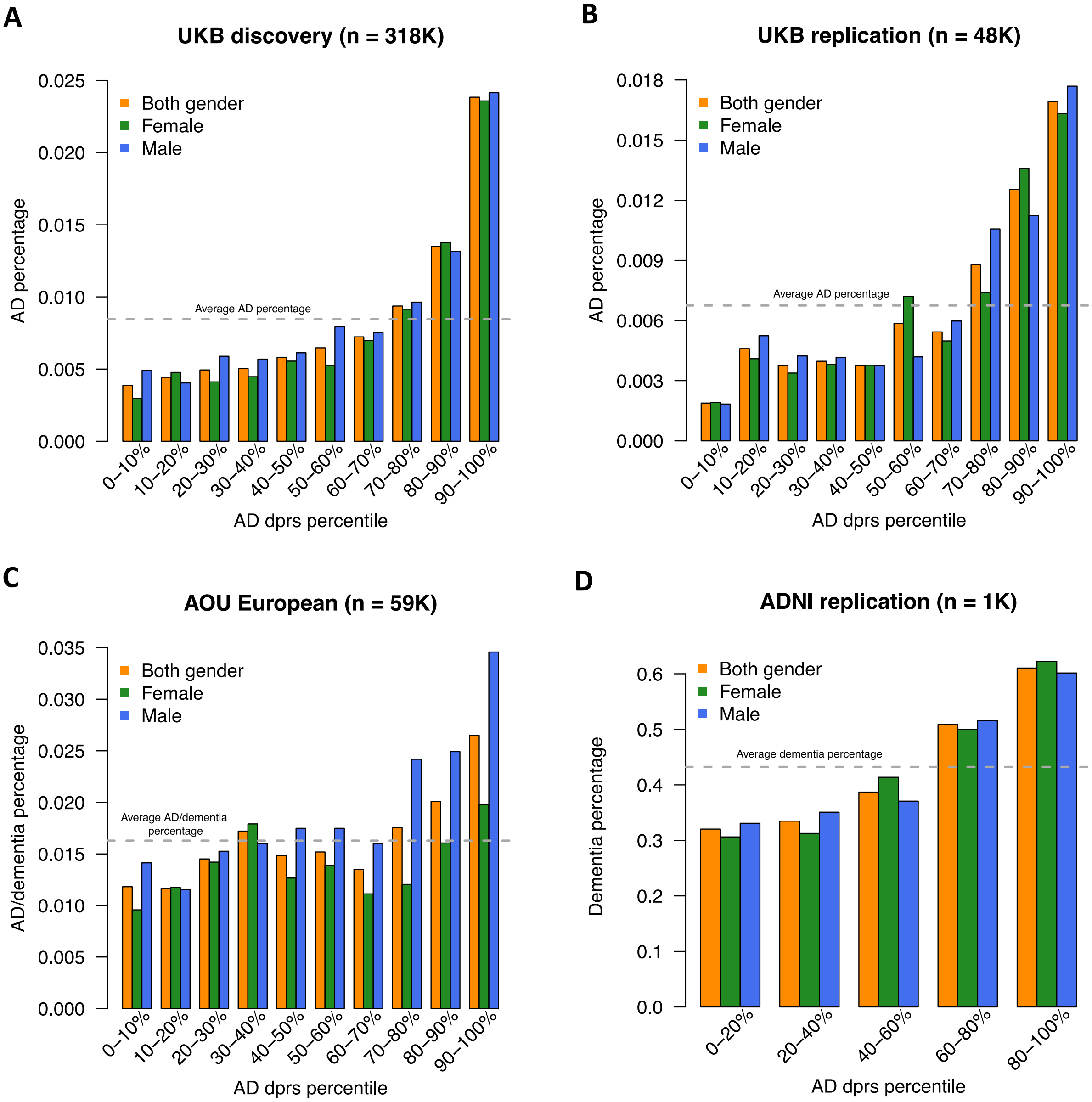


**Fig. S13 Alzheimer's disease dPRS analyses.**

**(A)-(D)** The disease percentage in different quantile groups of Alzheimer's disease (AD) dPRS, in the UKB discovery cohort, the UKB replication cohort, the AOU European cohort, and the ADNI cohort, respectively. The horizontal grey dashed line represents the disease percentage for the entire cohort.

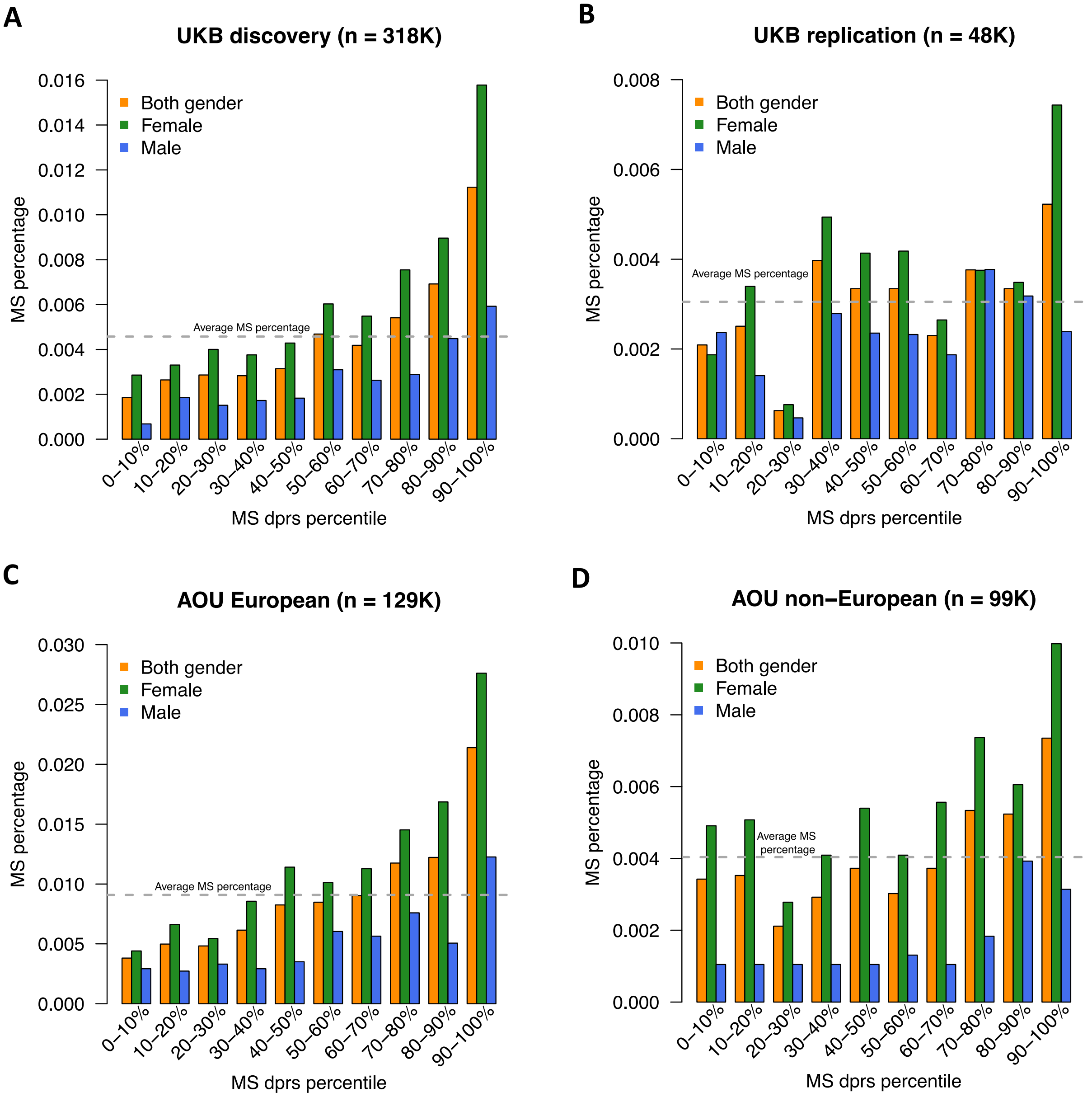


**Fig. S14 Multiple sclerosis dPRS analyses.**

**(A)-(D)** The disease percentage in different quantile groups of multiple sclerosis (MS) dPRS, in the UKB discovery cohort, the UKB replication cohort, the AOU European cohort, and the AOU non-European cohort, respectively. The horizontal grey dashed line represents the disease percentage for the entire cohort.

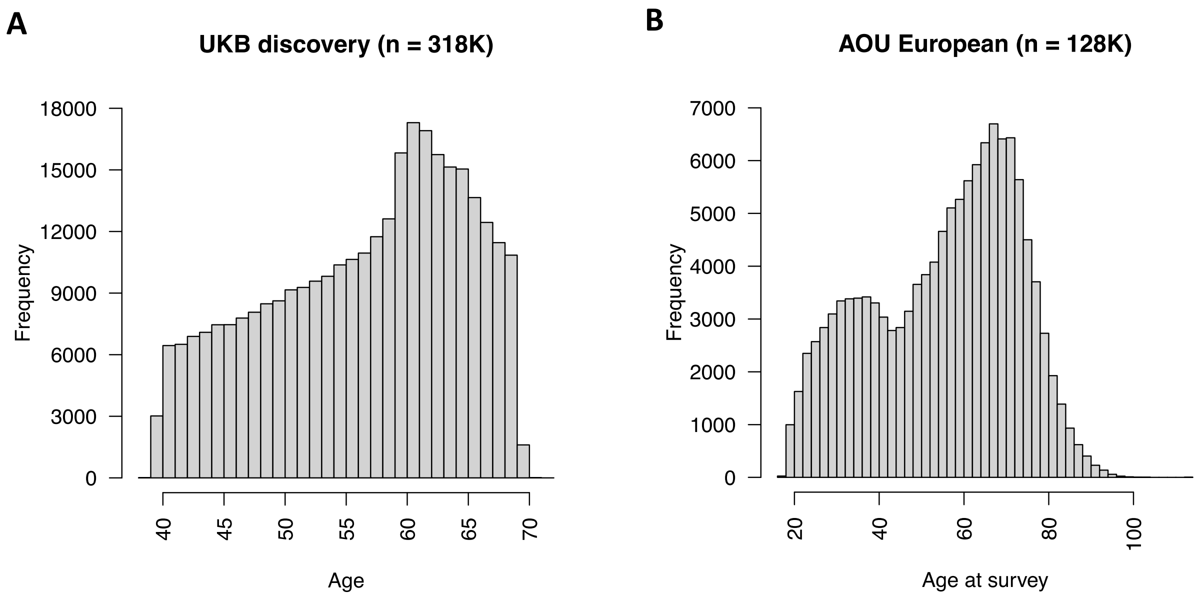


**Fig. S15 Age of participants in UKB and AOU.**

Age of UKB non-imaging participants and AOU participants.

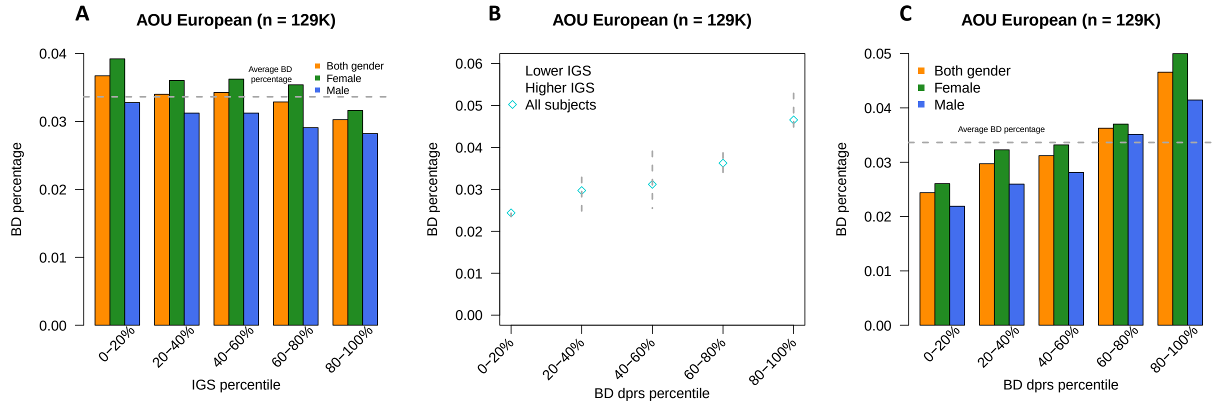


**Fig. S16 Bipolar disorder stratification on AOU.**

**(A)** For the pair between bipolar disorder (BD) and total.brain.volume (total brain volume), we show the disease percentage (*y*-axis) in quantile-based groups defined by total.brain.volume IGS (*x*-axis) in the AOU European cohort. **(B)** For each dPRS quantile group (*x*-axis), we show the BD percentage (*y*-axis) for all participants ("All subjects"), participants in the low 20% tail of total.brain.volume IGS ("Lower IGS"), and participants in the high 20% tail of total.brain.volume IGS ("Higher IGS"). **(C)** The disease percentage in different quantile groups of BD dPRS in the AOU European cohort. The horizontal grey dashed line represents the disease percentage for the entire cohort.

**Legends for Tables S1 to S18** (All tables can be found in a zip file).

Table S1. ID of brain and body imaging traits with corresponding IGS constructed.

Table S2. Performance of IGS of the 4,375 brain and body imaging traits on UKB European test set.

Table S3. Performance of IGS of the 4,375 brain and body imaging traits on UKB Asian test set.

Table S4. Performance of IGS of the 4,375 brain and body imaging traits on UKB African test set.

Table S5. ID of diseases used in IGS-disease stratification analysis.

Table S6. ID of phenotypes used in IGS-phenotype association analysis.

Table S7. Bonferroni-significant IGS-phenotype associations between phenotypes and IGS of BIG-KP brain MRI traits.

Table S8. Bonferroni-significant IGS-phenotype associations between phenotypes and IGS of UKB-Oxford brain MRI traits.

Table S9. Bonferroni-significant IGS-phenotype associations between phenotypes and IGS of body imaging traits.

Table S10. FDR-significant and replicated IGS-disease stratification results between IGS of selected brain and body IGS and diseases of the corresponding organ.

Table S11. FDR-significant and replicated IGS-disease stratification results between non-brain IGS and brain disorder, and between non-heart IGS and circulatory system diseases.

Table S12. AUC gain relative to the baseline model by including stratifying IGS in FDR-significant and replicated IGS-disease stratification results that are between IGS of selected brain and body IGS and diseases of the corresponding organ.

Table S13. AUC gain relative to the baseline model by including stratifying IGS in FDR-significant and replicated IGS-disease stratification results that are between non-brain IGS and brain disorder, and between non-heart IGS and circulatory system diseases.

Table S14. Mapping between phecode-based diseases that had FDR-significant and replicated IGS stratifications, and corresponding FinnGen R9 disease endpoints.

Table S15. AUC gain relative to the baseline-dPRS model by including the most stratifying IGS or all stratifying IGS in FDR-significant and replicated IGS-disease stratification results.

Table S16. Stratification results of Alzheimer's disease (AD) using selected FDR-significant brain IGS, age, AD dPRS and *APOE*.

Table S17. Stratification results of multiple sclerosis (MS) using selected FDR-significant brain IGS and MS dPRS.

Table S18. Stratification results of bipolar disorder (BD) using selected FDR-significant brain IGS and BD dPRS.
